# On the detectability and accuracy of computational measurements of enlarged perivascular spaces from magnetic resonance images

**DOI:** 10.1101/2023.07.25.23293140

**Authors:** Roberto Duarte Coello, Maria del C. Valdés Hernández, Jaco J.M. Zwanenburg, Moniek van der Velden, Hugo J. Kuijf, Alberto De Luca, José Bernal Moyano, Lucia Ballerini, Francesca M. Chappell, Rosalind Brown, Geert Jan Biessels, Joanna M. Wardlaw

## Abstract

Magnetic Resonance Imaging (MRI) visible perivascular spaces (PVS) have been associated with age, decline in cognitive abilities, interrupted sleep, and markers of small vessel disease. Therefore, several computational methods have been developed for their assessment from brain MRI. But the limits of validity of these methods under various spatial resolutions, and the accuracy in detecting and measuring the dimensions of these structures have not been established. We use a digital reference object (DRO) previously developed for this purpose, to construct an *in-silico* phantom for answering these questions; and validate it using a physical phantom. Our *in-silico* and physical phantoms use cylinders of different sizes as models for PVS. Using both phantoms, we also evaluate the influence of the “PVS” orientation on the accuracy of the diameter measured, different sets of parameters for two vesselness filters that have been used for enhancing tubular structures, namely Frangi and RORPO filters, and the influence of the vesselness filter *per-se* in the accuracy of the measurements. Our experiments indicate that PVS measurements in MRI are only a proxy of their true dimensions, as the boundaries of their representation are consistently overestimated. The success in the use of the Frangi filter for this task relies on a careful tuning of several parameters. The combination of parameters α=0.5, β=0.5 and c=500 proved to yield the best results. RORPO, on the contrary, does not have these requirements, and allows detecting smaller cylinders in their entirety more consistently in the ideal scenarios tested. The segmentation of the cylinders using the Frangi filter seems to be best suited for voxel sizes equal or larger than 0.4 mm-isotropic and cylinders larger than 1 mm diameter and 2 mm length. “PVS” orientation did not influence their measures for image data with isotropic voxel size. Further evaluation of the emerging deep-learning methods is still required, and these results should be tested in “real” world data across several diseases.

## Introduction

Perivascular spaces (PVS), eponymously named Virchow-Robin spaces after the 19^th^-century anatomists and pathologists Rudolf Virchow and Charles Philippe Robin, are fluid-filled spaces that surround the walls of the blood vessels. PVS in the brain have attracted the attention from the scientific community due to their suspected role in waste clearance and thus their potential value as an imaging biomarker of brain health function (Wardlaw et al., 2020; Donahue et al., 2021; Sepehrband et al., 2021). Their increase in size to diameters from 1 to 3 mm and lengths from 3 to 5 mm (Valdés Hernández et al., 2013), prompting their visibility in magnetic resonance images (MRI), has been associated with cognitive dysfunction in the elderly (Hilal et al., 2018; Passiak et al., 2019) possibly even more strongly than other markers of small vessel disease (Passiak et al., 2019), sleep dysfunction (Aribisala et al., 2020; Baril et al., 2022; Berezuk et al., 2015; Del Brutto et al, 2019), inflammatory markers (Aribisala et al., 2014), hypertension (Dubost et al., 2019), ageing, brain lacunes, and microbleeds (Francis et al., 2019). PVS burden has also been associated with the speed of white matter deterioration in older adults, and partially mediating the effect of sleep in white matter health (Aribisala et al., 2023). It is therefore not surprising that several methods for assessing PVS from magnetic resonance images (MRI) have been developed, ranging from visual scoring systems (Patankar et al., 2005; Potter et al., 2015) to fully-automated computational approaches (Pham et al., 2022; Barisano et al., 2022).

While neuroradiological scoring has been largely considered the reference standard, advances in scientific and research methodologies have made large-scale research a norm. Large databases that combine clinical, demographic, genetic and imaging data are rapidly emerging to underpin and advance clinical research, but they are only partially annotated, and expert visual assessment at such scale is implausible. Also, due to the nature (i.e., void spaces once dissected) and size of this features (i.e., wide range in microscopic scale) and post-mortem tissue collapse, comparison of *in-vivo* MRI measurements with histology will be imprecise. Automatic methods to assess brain enlarged PVS would enable analyses of very large studies. However, they have been evaluated against neuroradiological ratings and manually-annotated images in only one slice per region of interest or in only few cases. Moreover, the limits of validity of these techniques and accuracy levels in measuring PVS are still not known. MRI from the human brain alone is unlikely to provide this information as measurements of small brain structures are compromised by image resolution, confounding pathologies, and artefactual effects, the CSF pulsation due to the heartbeat being one of them. Previous study presented a Digital Reference Object (DRO) designed for this purpose (Bernal et al., 2022) and evaluated the performance of three image enhancement methods under various spatiotemporal imaging considerations including sampling, motion artefacts, and Rician noise. We here use this previously developed DRO to develop an *in-silico* phantom, and use it together with a physical phantom to inform on a) the influence of the choice of filter parameters in the measurements b) how the spatial resolution of the image influences the PVS diameter measured, c) the influence of PVS orientation in the accuracy of the measurements, d) the differences introduced by the choice of filtering methods, and e) the detectability of PVS in research protocols using MRI scans with 1 mm^3^ isotropic voxels.

## Materials and Methods

### PVS DRO

PVS are seen in MRI as thin linear or small punctate structures depending on the visualisation axis, with signal intensity similar to that of CSF (Valdés Hernández et al., 2013; Wardlaw et al., 2013). Using this description, Bernal et al. (2022) choose a cylinder as a geometric model of PVS. The parametric equation of a cylinder is given by:

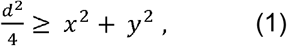

for -*l*/2 ≤ *z* ≤ *l*/2, where x, y and z are the 3D coordinates, *l* is the cylinder length and *d* is the diameter of the cylinder. The volume of the cylinder is calculated using

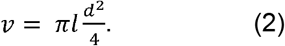

### In-silico phantom

We define our PVS binary mask (1: if the voxel is inside the cylinder; and 0: otherwise) using equation (1) and consider it a PVS representation in high resolution. We, then, reduce the resolution using a linear interpolation to simulate PVS in a lower-resolved image and introduce partial volume effects. The intensity of the voxel in a given coordinate (x,y,z) of the image space is then modelled via the following equation:

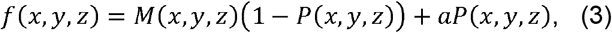

where *M* is the intensity of the background (usually white matter intensity), *P* is the proportion of PVS in the voxel (i.e., the full PVS “body” or a border with partial volume), and *a* is the intensity of the CSF in the scan (i.e., equal to the intensity of the simulated PVS).

We generate several “PVS” in our *in-silico* phantom by varying the diameter, length and orientation of the cylinders. In particular, we vary the diameter from 0.2 to 3 mm, the length from 1.02 to 13 mm, and we rotate the cylinders around the x-axis from 0° to 180° and the z-axis from -45° to 45°. We confine each cylinder within a space of 15 × 15 × 15 mm^3^ as per maximum length definition (Valdés Hernández et al., 2013; Wardlaw et al., 2013), and to avoid them being too close to each other. We constructed our *in-silico* PVS phantom by placing together all the cubes of 15 × 15 × 15 mm^3^, as can be seen in Figure 2.

**Figure 1.**
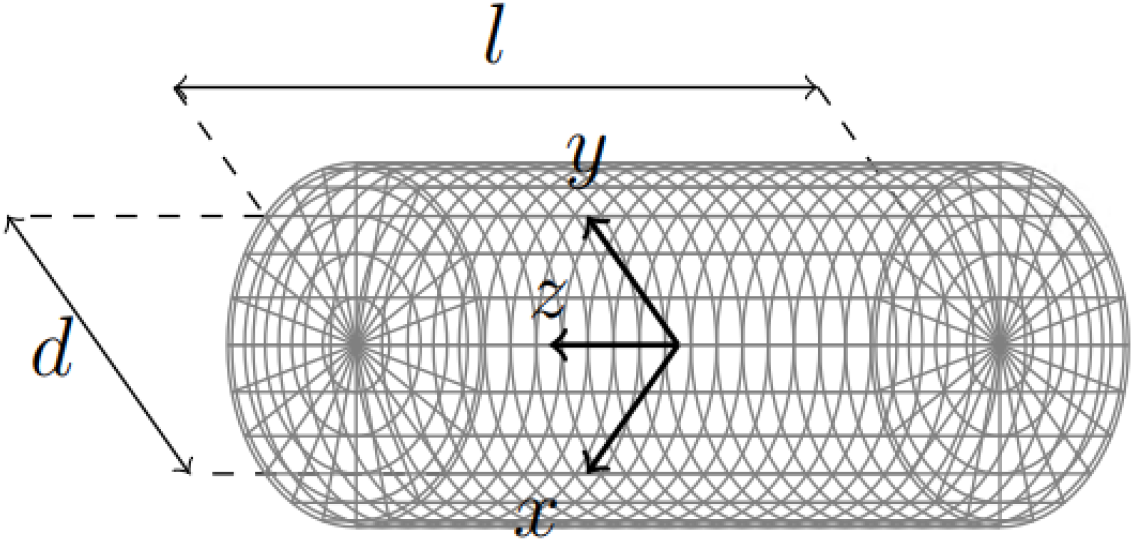
Cylinder representing a linear PVS or a linear section of it, defined by equation (1) with length *l* and diameter *d*. The origin of the 3D space coordinates (*x, y, z*) to place this digital reference object is at the object’s centre.

**Figure 2.**
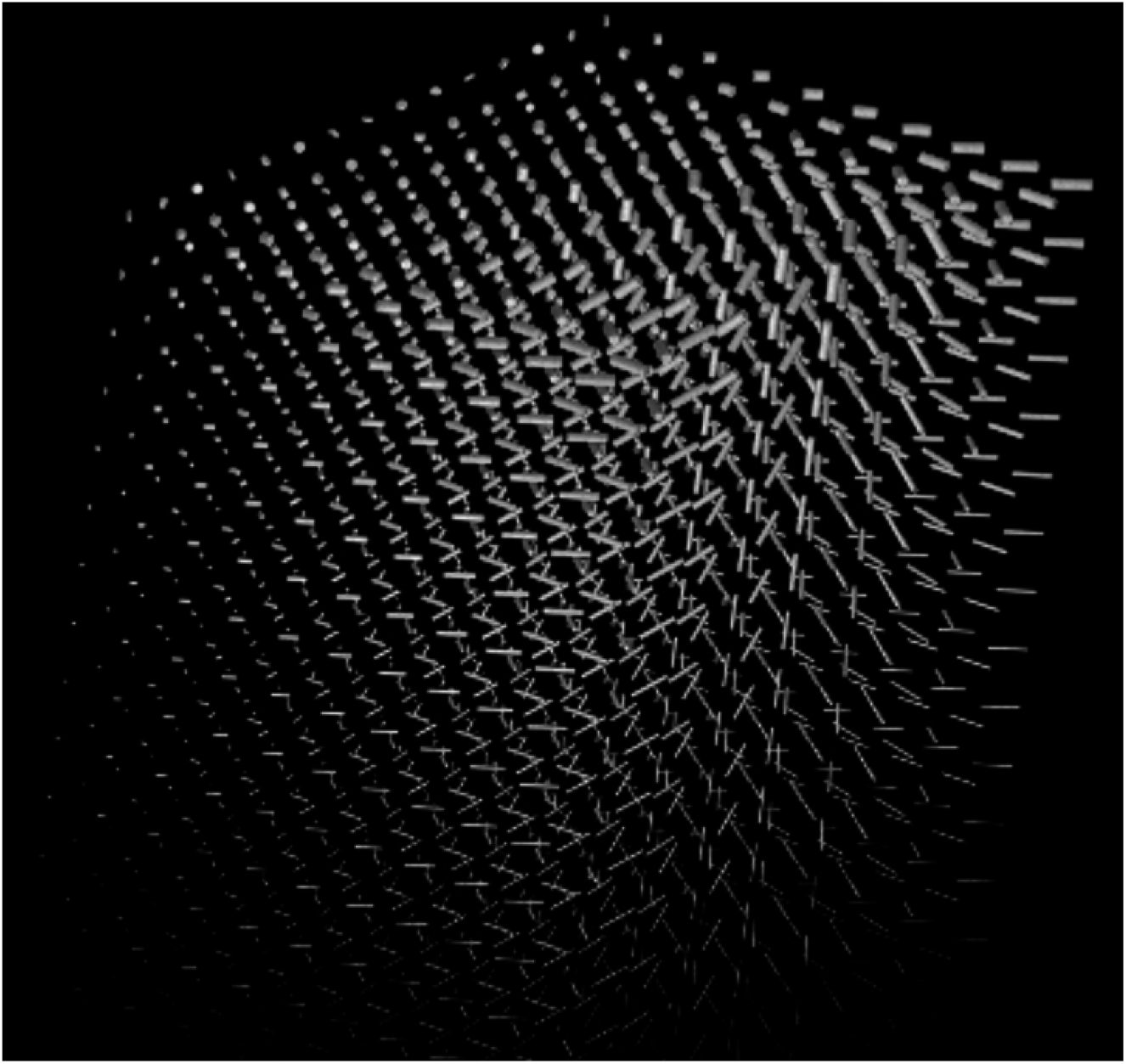
*In-silico* PVS phantom constructed by placing together cubes of 15 × 15 × 15 mm^3^, each containing a cylinder (i.e., DRO) with different widths, lengths and orientations.

### UMC Utrecht PVS phantom

We analysed images from a physical phantom built at the Utrecht University Medical Center with the primary purpose of investigating the influence of spatial resolution, orientation, and image analysis method, on the accuracy of measurements of PVS in a 7T MRI scan, in parallel to validate our *in-silico* phantom. The UMC Utrecht PVS phantom consists of a Perspex plate with holes of diameters ranging from 0.2 to 3 mm, filled with water, and scanned with a T2-weighted contrast using a turbo spin-echo sequence in a 7T MRI scanner (Figure 4). The acquisition was similar to the acquisition used in previous work on imaging PVS in human volunteers (Bouvy et al., 2014), but slightly adjusted to the small phantom size, and adjusted to yield protocols for multiple isotropic resolutions. See Supplementary Table S1 for an overview of the scan parameters. Subsequently in this paper, we will mention the reconstructed voxel sizes while referring to the resolution of the images unless stated otherwise to facilitate the analyses and interpretation of the results. These are larger than the acquired voxel sizes. The acquisition resolution that corresponds to each of the resolutions of the reconstructed images can be seen in Supplementary Table S1. In addition to the default orientations (i.e., coronal, sagittal and transverse), the field-of-view was also rotated 15°, 30°, and 45° around the ‘feet-head’ axis, which yields the most unfavourable positioning (i.e. these are rotations around an axis that is perpendicular to the centerlines of the holes).

**Figure 3.**
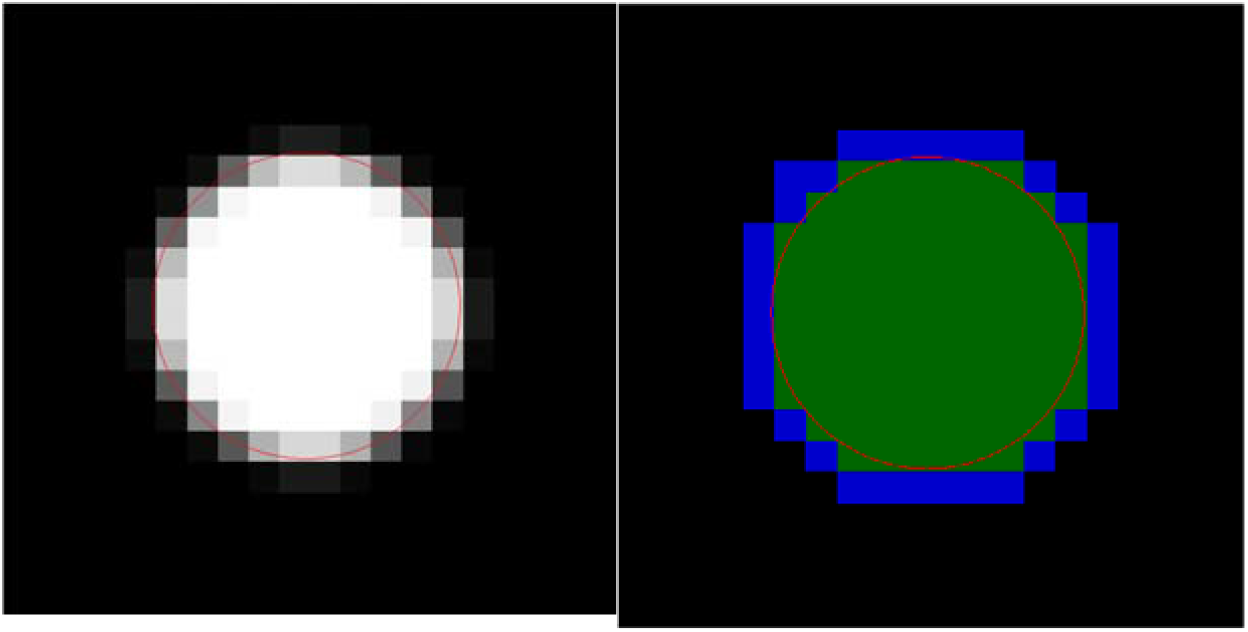
Transversal cut of an example of PVS DRO with diameter 3 mm in isotropic voxels of 0.3 × 0.3 × 0.3 mm^3^. (Left) Intensity image (Right) Segmentation mask. The red circle (i.e., thin perimeter line in the intensity image) indicates the true boundaries of the DRO, the green voxels are the voxels where the DRO occupies at least 30% of the voxels and the blue voxels are the surface of the DRO. The boundaries of the PVS representation, i.e., the measured dimensions, may be overestimated due to the partial volume effect (Figure 3), as per Bernal et al. (2022).

**Figure 4.**
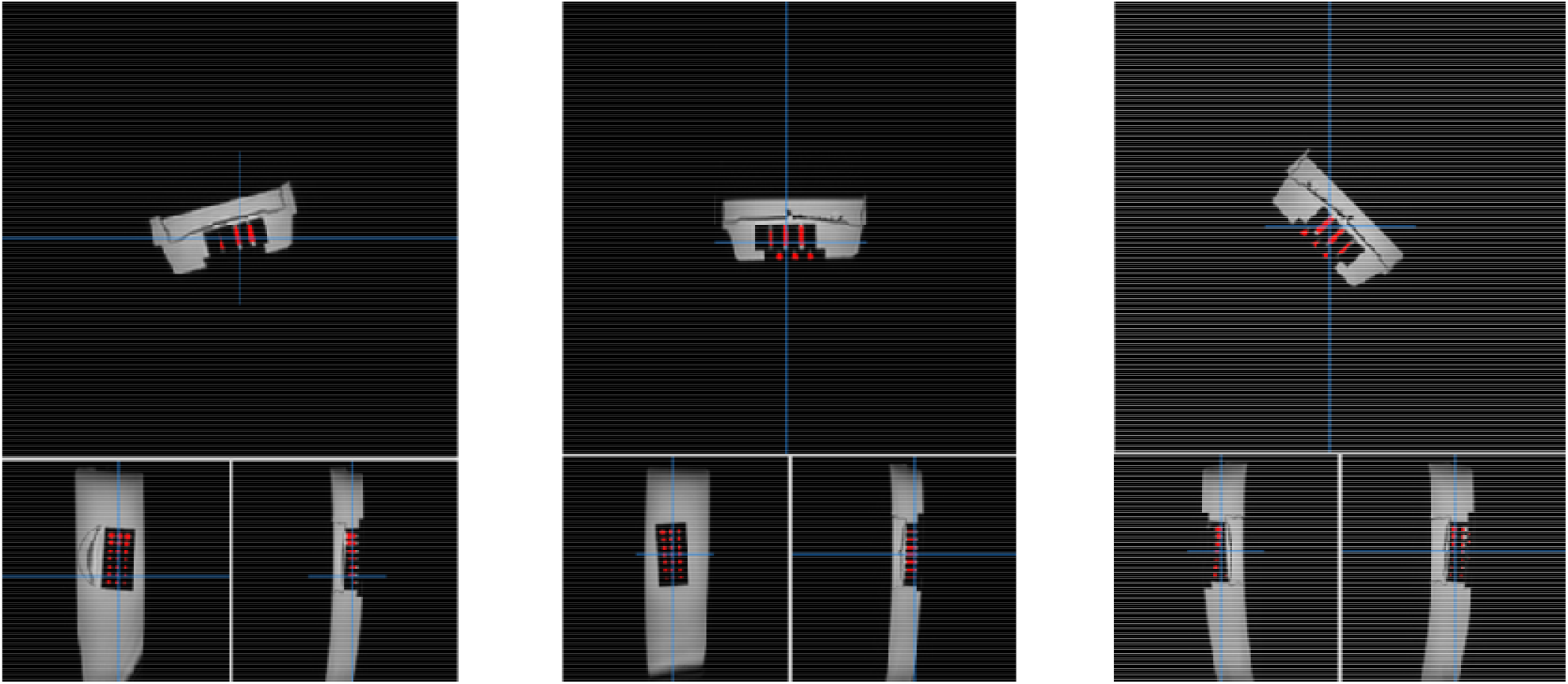
UMC Utrecht physical phantom scanned with the field-of-view rotated at different angles. (Left) 15 degrees, (Centre) 0 degrees, (Right) 45 degrees.

Figure 5 shows the images obtained. Holes of 0.2 and 0.3 mm diameters could not be filled with water. Therefore, they are not visible. Holes with diameters smaller than the voxel size are visible but attenuated due to partial volume effect. The PVS measured diameter in a scan depends on the location of the PVS relative to the voxel. Ideally, a PVS would run exactly through several voxels, but often it is located on the edge of two or various voxels or pass diagonally through a set of voxels. Here the holes representing PVS can appear enlarged as usually they are not aligned to the centre of the voxels they cross. The oblique length in the plane (𝓁2D) and the length of the body diagonal (𝓁3D) can be calculated as the square root of the sum of squares of the voxel dimensions (i.e., 𝓁2D = sqrt(x^2^ + x^2^) and 𝓁3D = sqrt(x^2^ + x^2^ + x^2^) for isotropic voxels of dimension x). In such way, for different settings, the lengths of the holes representing PVS can differ (see Supplementary Table S2).

**Figure 5.**
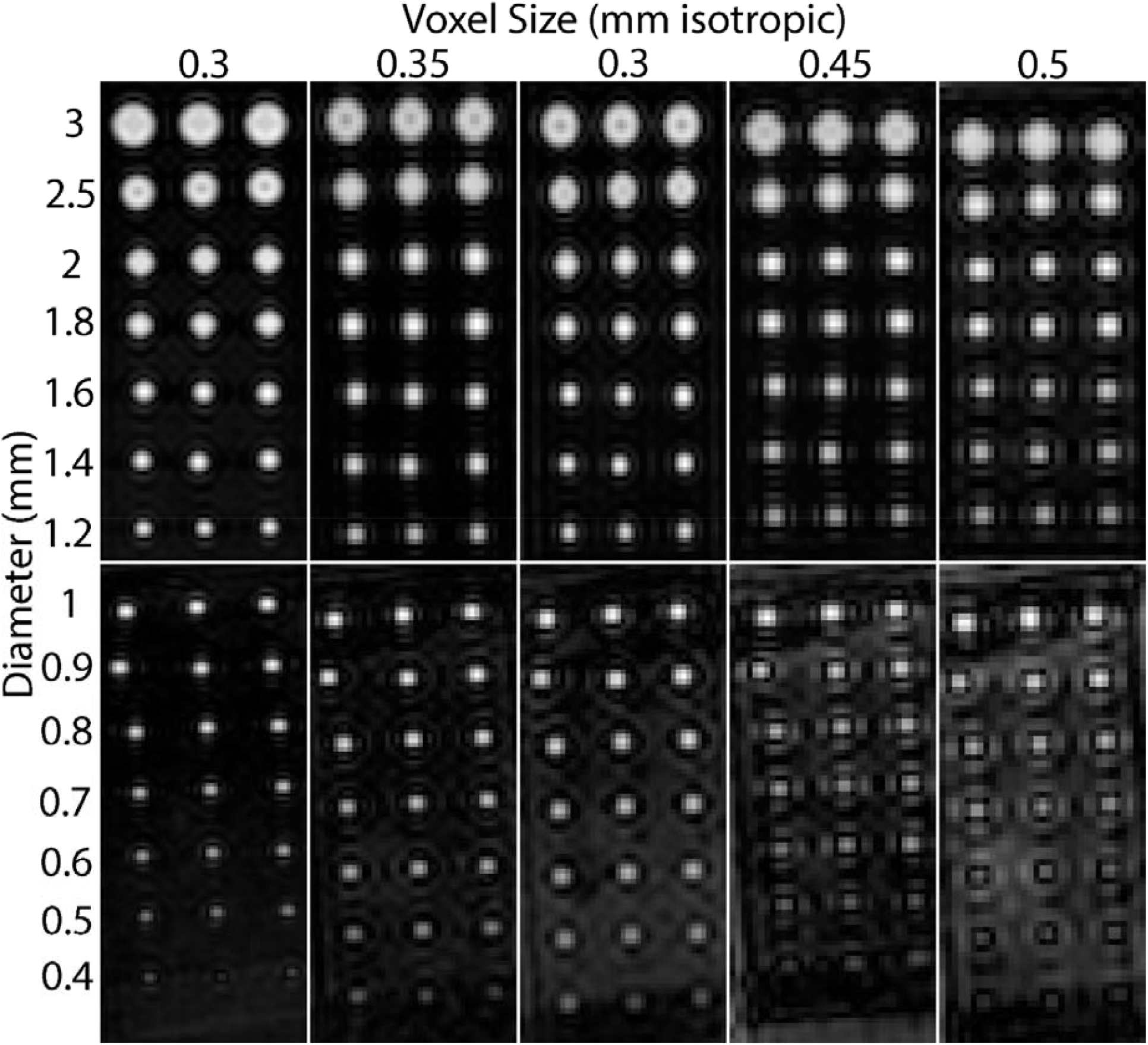
Transversal slices from the UMC Utrecht phantom acquired with different resolutions. From left to right, reconstructed voxel sizes: 0.3, 0.35, 0.4, 0.45 and 0.5 mm-isotropic. (the acquired “physical” resolution was 0.6, 0.7, 0.8, 0.9 and 1.0 mm isotropic, respectively, see Suppl. Table S1). From top to bottom in upper panels hole diameters 3, 2.5, 2, 1.8, 1.6 and 1.2 mm; and 1, 0.9, 0.8, 0.7, 0.6, 0.5 and 0.4 mm in bottom panels. Please note that *k*-space ringing artefacts are more apparent with larger voxel sizes.

### Automatic PVS segmentation

We evaluate the performance of one of the fully-automated image processing/analysis methods most widely used in clinical research at present for segmenting PVS, which consists of enhancing PVS-like structures using a “vesselness” filter, thresholding the resulting “vesselness” likelihood map, and analysing the morphology of the segmented PVS. We evaluate two “vesselness” filtering methods, which are representative of the two classes of filters mostly used: 1) Hessian-based filters that rely on information provided by the Hessian matrix of the image, and 2) the morphological-based filters that rely on the mathematical morphology of the image intensities. We use the Hessian-based Frangi filter (Frangi et al., 1998), firstly used for PVS segmentation by Ballerini, et al. (2018), and the morphological-based Ranking Orientation Responses of Path Operators (RORPO) (Merveille et al., 2018), firstly used for PVS segmentation by Bernal et al. (2022).

We explore the influence of the filter parameters in the segmentation, and further analyse the limits of validity and expectations of accuracy in the measurements of PVS-like structures using the optimal thresholds and the parameters previously published after these being validated in the corresponding pioneer publications. As such, for these analyses, the parameters of the Frangi filter were set as suggested in (Ballerini, et al., 2016, 2018) *α* = 0.5, *β* = 0.5 and c = 500, as we show in the analyses of the parameters of the Frangi filter that these values yield the best results. The scales of each filter were tuned for the image resolution. The intensities of each image were normalised to values from 0 to 255 only considering the region of interest (ROI). The filters were also applied to the images from the UMC Utrecht physical phantom. The optimum threshold applied to the resultant “vesselness” map was the one which allowed capturing holes across the whole (or most of the) range of diameters, and it was selected only on the image with the highest resolution and applied to the rest. The resulting segmentation mask was used to compute the results. Please note that we chose the threshold based on the physically acquired scans, because they have noise and artefacts that are not reflected in our *in-silico* phantom.

### Image analyses

Using the *in-silico* phantom, we simulated the UMC Utrecht PVS (physical) phantom with voxel sizes varying from 0.3 to 0.5 mm (Supplementary Figure S1), and calculated the diameters of the cylinders in both phantoms using the MATLAB built-in function regionprops3. We plotted the ‘true’ segmentation and the results obtained from each filtering method for each phantom (i.e., *in-silico* and physical) to illustrate the expectations in accuracy of the PVS segmentation under ideal conditions (i.e., in absence of noise and confounding artefacts) for each spatial resolution, and to evaluate the fidelity with which our *in-silico* phantom can be used to represent a physical MRI phantom for evaluating the accuracy of PVS segmentations. We also analysed the impact of PVS orientation in the measurement accuracy at voxel sizes of 0.35 × 0.35 × 0.35 mm^3^ in both phantoms. We further analysed the limits of validity of the computational measurements that use the two filtering methods evaluated, in images with 1mm-isotropic voxels, as this is the spatial resolution commonly used in medical research imaging protocols. For this, in our *in-silico* phantom, we generated cylinders from 0.1-3 mm diameter and from 0.1-10 mm length with different orientations and plotted the results. If at least 1 voxel inside a cylinder could be identified, then it was considered as detected. Otherwise, it was considered as not detected.

### Statistical analyses

Differences between all the cylinder diameters’ measurements at each voxel size were investigated using box plots. The average measurements, range, and absolute differences from the real (i.e., ideal) measurements of the cylinders obtained from using each filtering method were compared using the paired Wilcoxon sign rank test. Bland-Altman analyses of the diameters’ measurements were also performed, but excluding the undetected cylinders (i.e., those for which the measurement diameter equalled zero).

## Data availability statement

The data and source code correspondent to the analyses contained in this manuscript are publicly available from https://doi.org/10.7488/ds/7454.

## Results

### Influence of the Frangi filter parameters

Supplementary figures S4 to S9 illustrate the effect of different values of α, β, c and scale parameters of the Frangi filter for the identification of the PVS-like structures. A spatial resolution (i.e., voxel size) of 0.3 mm-isotropic was chosen to represent these effects. Alpha (α) controls the sensitivity of the filter in differentiating between tube-like and plate-like structures. Values of α that are too low allow more plate-like structures to be highlighted. As the length of the PVS of interest (i.e., the ones that are enlarged) ranges between 3 to 5 mm, α=0.5 was best suited to dismiss the smaller structures while increasing the sensitivity to distinguish elongated cylinders mimicking large PVS from wedge-like structures that could mimic typical lacunes (see results from α values of 0.1, 0.5 and 0.9 in Supplementary Figure S4). Beta (β) controls the sensitivity of the filter in differentiating between tube-like and blob-like structures. Lower values of β reduce the sensitivity in detecting short cylinders and the filter response in the extremes of larger cylinders (Supplementary Figure S5, left hand-side panel, β=0.1). Values of β that are too high, allow more blob-like structures to be highlighted and cause a blob-like effect in the extremes of the cylinders (Supplementary Figure S5, right hand-side panel, β=0.9). The parameter c controls the sensitivity of the contrast between the bright/dark object (PVS) and the background (commonly normal-appearing brain tissue), but lower values of c could also increase false positives (see effects of values 50, 500 and 100 in Supplementary Figure S6). The maximum scale parameter controls the sensitivity to the size of the cylinders. In principle, higher values of the maximum scale will detect bigger tubular objects (Supplementary Figure S7), but it depends on the voxel size as well. Smaller voxel sizes require higher values compared with larger voxels to detect tubular structures of the same length. The scale ratio controls the sensitivity to detect cylinders of certain aspects’ ratios, complementing the maximum scale parameter, but at a cost in computational time. Finer scales detect better the tubular structures but increase the computational time. Coarse scales would reduce the computational time but at the cost of not detecting some of these structures (Supplementary Figure S8). Also, suboptimal scales can lead to either under or over-estimation of the size of the PVS. If there is over-estimation we could end up merging PVS that are close together altering the real count.

### Influence of RORPO filter parameters

The RORPO filter only uses one parameter: the maximum scale representing the maximum length of the tubular objects to identify. Supplementary Figure S9 illustrates the effect of this parameter for a spatial resolution of 0.3 mm-isotropic. In principle, higher values of the maximum scale will detect bigger tubular objects (Supplementary Figure S9). The filter seems to be more sensitive to objects that are aligned to the Cartesian coordinates. The filter output inside the cylinders is more homogeneous than the output of the Frangi filter, thus explaining the more linear behaviour in Figure 6.

**Figure 6.**
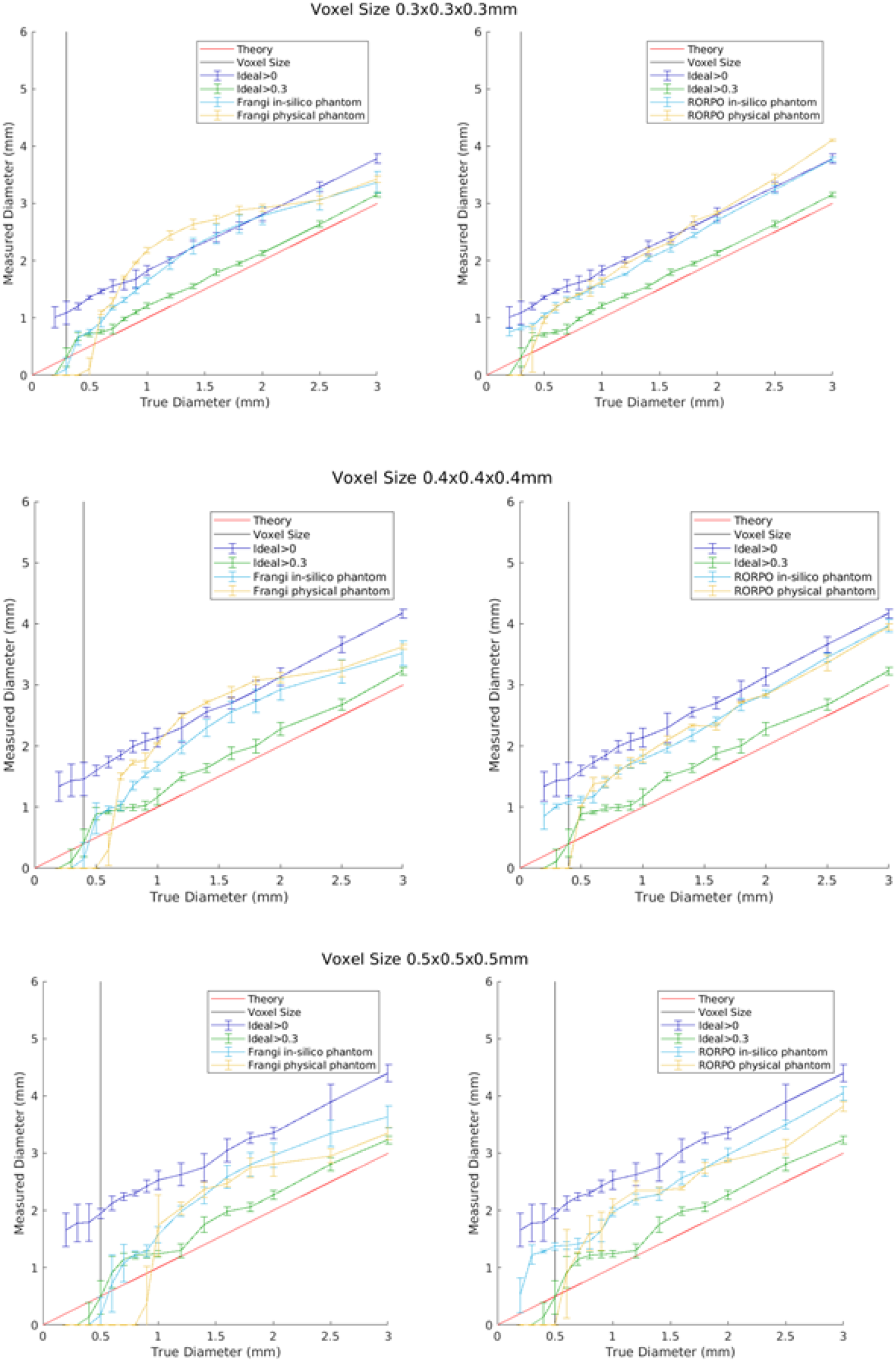
Measurements of cylinder’s diameters using the Frangi (left hand-side graphs) and RORPO (right hand-side graphs) filters for their detection in both phantoms, with isotropic voxel sizes of 0.3, 0.4 and 0.5 mm-isotropic and acquired resolutions equal to twice the reconstructed voxel sizes. The red line corresponds to the theoretical diameters, the black line corresonds to the voxel size of the images, the dark blue line corresponds to the ideal segmentation of the *in-silico* phantom considering all voxels of the cylinders, the green line corresponds to the ideal segmentation of the in-silico phantom considering voxels located at least 30% inside the cylinders, the light blue line corresponds to the results for the in-silico phantom and the yellow line corresponds to the filter results for the physical phantom.

### Accuracy of the PVS measurements at different spatial resolutions

Figure 6 shows the results from measuring the diameter of the cylinders of both phantoms at spatial voxel sizes of 0.3, 0.4 and 0.5 mm-isotropic, and Figure 7 shows the Bland-Altman plots of the differences between the average diameter measurements from both filters and the ideal cylinder diameters at voxel sizes of 0.3, 0.35, 0.4, 0.45, and 0.5 mm-isotropic (see also Supplementary Figure S2). The RORPO filter had increased sensitivity compared to the Frangi filter for detecting small cylinders, between 0.5 and 2.0 mm diameters regardless of the voxel size, while the Frangi filter yielded better results when measuring diameters higher than 1.0 mm for voxels equal or bigger than 0.45 mm-isotropic (i.e., from the ones evaluated in these experiments) (See Supplementary Table S5). Measurements obtained using RORPO showed a linear correspondence between the measured diameter and the real diameter (i.e., ideal value) across the range of diameters evaluated. For smaller voxel sizes (i.e., 0.3 and 0.4 mm-isotropic), diameter measurements were closer to the ideal segmentation of the *in-silico* phantom considering all voxels of the cylinders (i.e., including those with partial volume effects), while for voxel sizes of 0.5 mm-isotropic, they were closer to the ideal segmentation of the in-silico phantom considering voxels located at least 30% inside the cylinders (e.g., green masked area in Figure 3 right panel). Deviations from the ideal measurements, e.g. non-uniformities of the filter output inside the objects, were also observed. The measured diameters in both (i.e., *in-silico* and physical) phantoms followed similar pattern across sizes when compared to the theoretical values, in addition of being close to each other, thus providing confidence in the use of our in-silico phantom model to establish the limits of validity of the PVS segmentation methods. As expected (see Figure 3 and Supplementary Table S2), almost all measurements were larger than the real dimensions of the cylinders (Figures 6 and 7).

**Figure 7.**
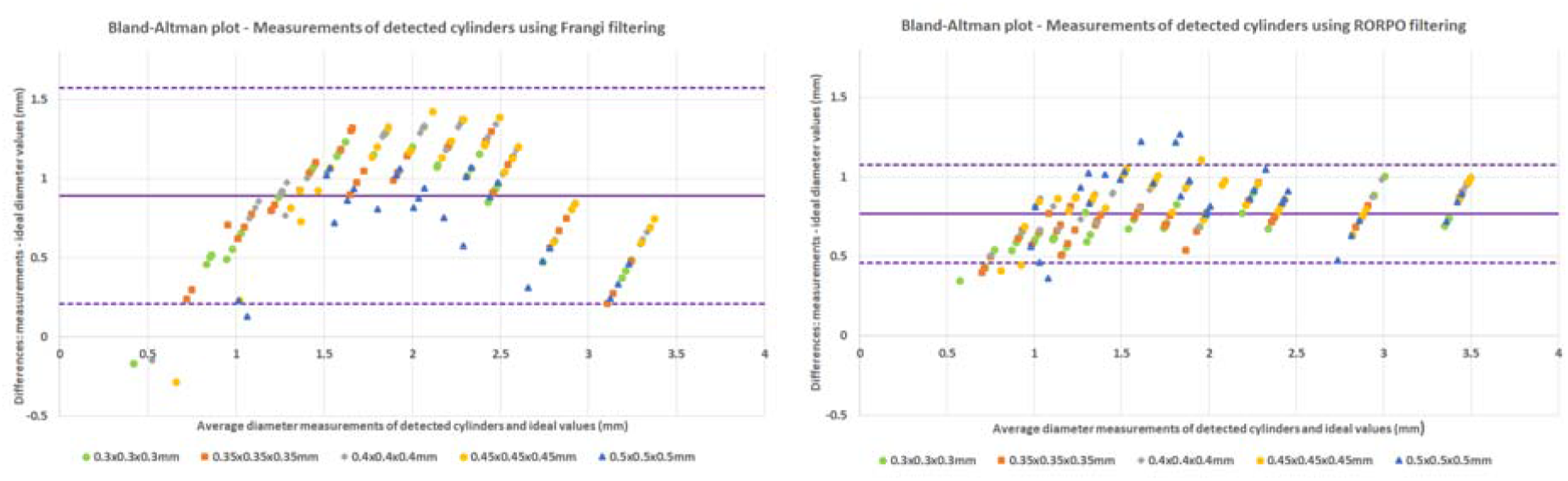
Bland-Altman plots of the differences between the valid measurements from Frangi (left) and RORPO (right) at different voxel sizes (VS) and the real diameter of the cylinders of the physical phantom. The scales of the diameters are in mm (i.e. horizontal and vertical axes), and identical in both graphs. Mean difference values for each voxel sizes are tabulated in Supplementary Table S5.

Supplementary Table S3 contains the values of the diameter measurements in mm using the Frangi filter, the filtering method that has been most used in clinical research (Pham et al., 2022), to identify the cylinders of the physical phantom. The mean absolute differences between the real diameters and the measured ones for voxels of 0.3 mm-isotropic was 0.76 (SD=0.37, range=[0.2, 1.24]) mm. For voxels of 0.35 mm-isotropic it was the same but with range extending up to 1.27 mm. For voxels of 0.4 mm-isotropic the mean absolute differences were the highest: 0.81, (SD=0.39, range=[0.2, 1.31]) mm, and then declined for voxels of 0.45 mm-isotropic (0.78 (SD=0.35, range=[0.2, 1.27]) mm), to be the smallest (in average) for voxels of 0.5 mm-isotropic (0.62 (SD=0.24, range=[0.2, 0.96]) mm).

### Effect of PVS orientation in measurements

Figure 8 illustrates the diameters measured only on coronal orientation (upper row), and combining the measurements obtained from quantifying the cylinders placed (i.e., in the *in-silico* phantom) or scanned (i.e., in the UMC Utretch physical phantom) in different orientations. Although even the real values slightly differ when the scan is acquired in a perpendicular or parallel plane to the cylinders with respect to those from when the images are acquired in oblique planes, these differences were very small if the same image processing method (i.e., filter) was used (see Supplementary Figure S3 and the dataset in https://doi.org/10.7488/ds/7454 for individual measurements and their average values in each orientation).

**Figure 8.**
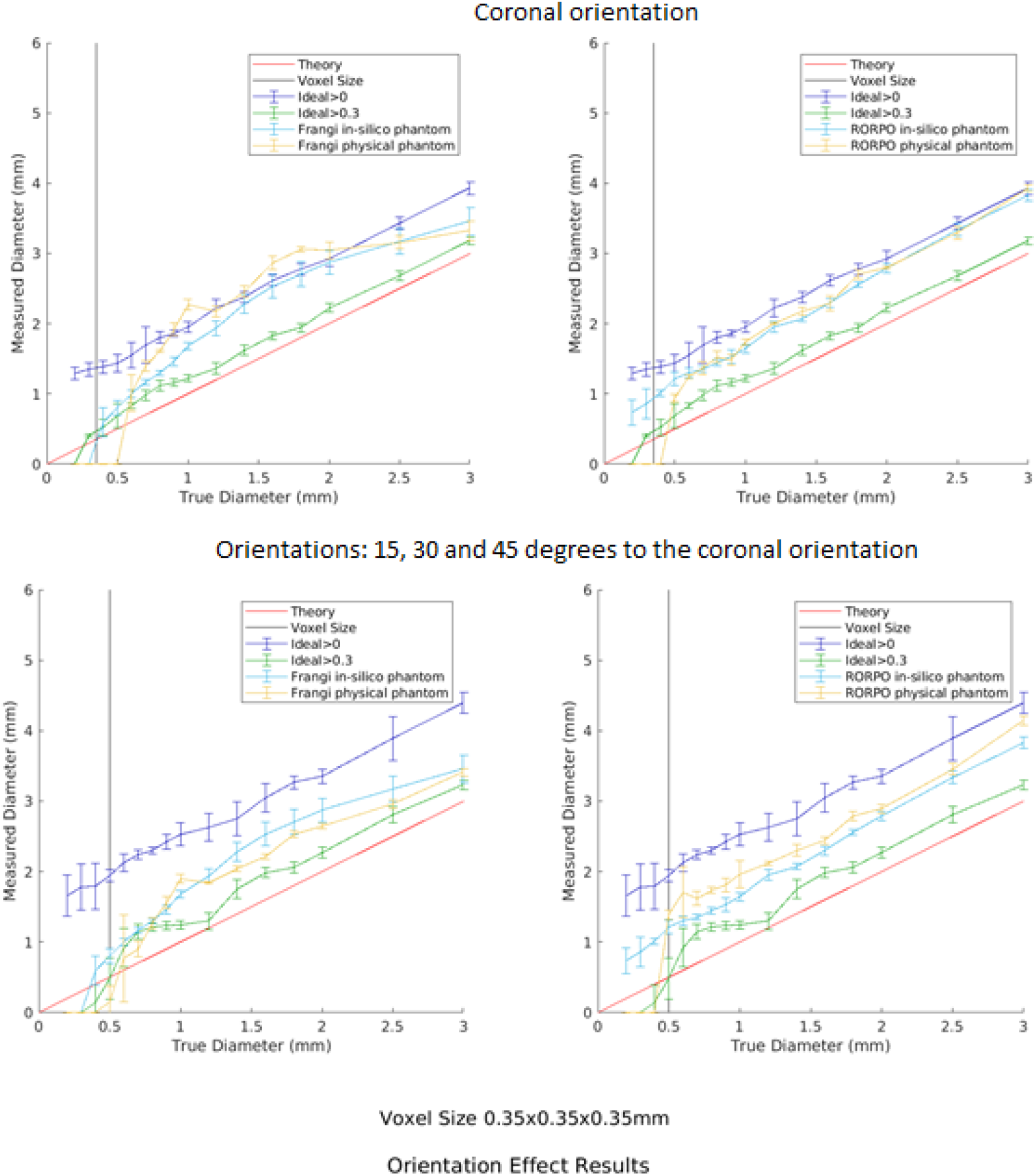
Results of measuring the cylinders’ (PVS-DROs’) diameters in the *in-silico* and physical phantoms only in coronal orientation (upper row) and averaging the diameters’ measurements from four different orientations: coronal, and 15°, 30° and 45° foot-head with respect to the coronal (bottom row) using the Frangi filter (left hand side plot) and RORPO (right hand side plot). The voxel size is 0.35 × 0.35 × 035 mm^3^ and the acquired resolution was equal to twice the reconstructed voxel size. The red line corresponds to the theoretical diameters, the black line corresonds to the voxel size of the scans, the dark blue line corresponds to the ideal segmentation of the PVS-DROs considering all their voxels, the green line corresponds to the ideal segmentation of the PVS-DROs considering voxels located at least 30% inside them, the light blue line corresponds to the results for the *in-silico* phantom, and the yellow line correspond to the filter results for the physical phantom.

### In-silico measurements in images with 1 mm^3^isotropic voxels

The limits in detecting cylinders of different diameters from both filters for voxels of 1 mm-isotropic can be seen in Figure 9. With the given filter parameters and thresholds, the Frangi filter could detect cylinders with at least 2 mm of length and 1 mm diameter. The RORPO filter could detect narrower cylinders of 0.4 mm diameter.

**Figure 9.**
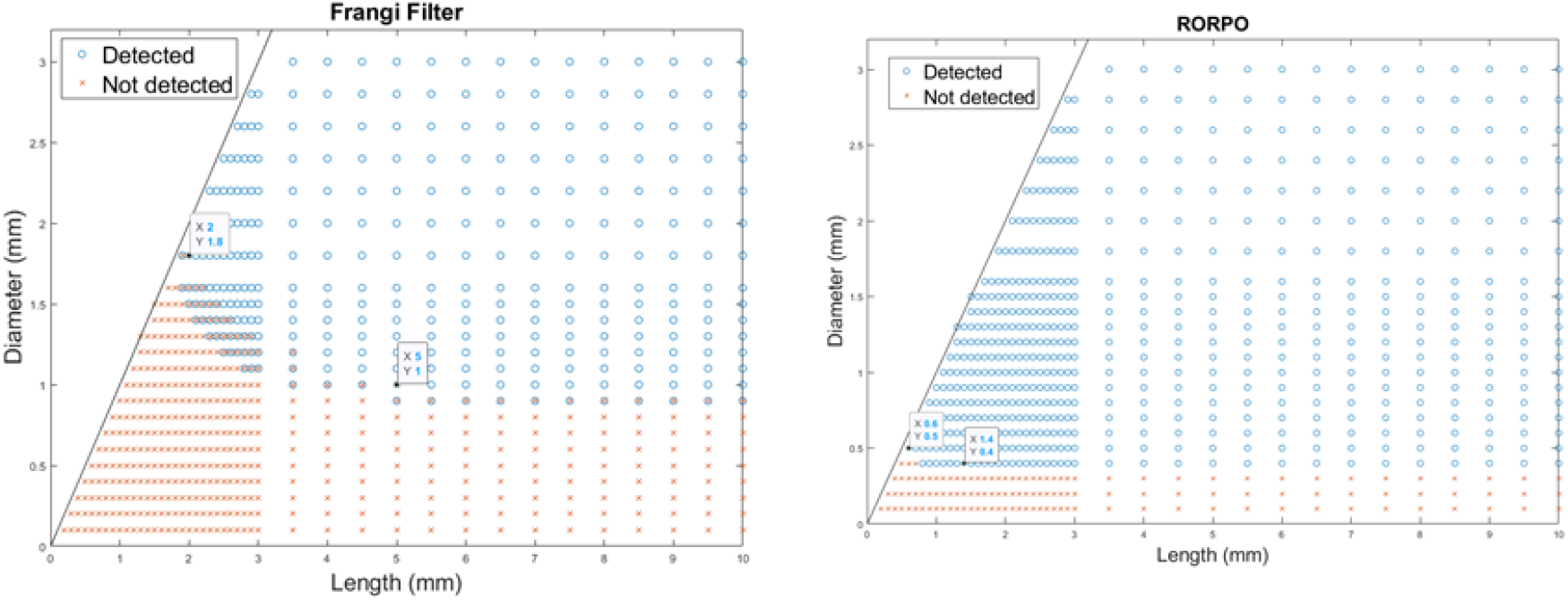
Results from applying the Frangi (left) and RORPO (right) filters. Cylinders with diameter and lengths represented by the red crosses were not detected, whereas the ones represented by the blue circles were detected. Overlap in detection/not detection across the range of dimensions in the graphs is owed to the different spatial orientations of the cylinders. The cylinders with diameter greater than their length do not follow the definition of PVS and were not considered in the experiment.

Figure 10 shows the length and diameter obtained from applying each filter with their optimal parameters. Cylinders (i.e., PVS-DROs) that did not have the minimum dimensions for being detected by each filter (as per results shown in Figure 8) were removed from this analysis. Measurements with the Frangi filter were closer to the ideal segmentation of the in-silico phantom considering only voxels with at least 30% inside the cylinder, and overlapped with these ideal segmentations in length. Measurements using RORPO were closer to the ideal segmentation considering all voxels (i.e., either fully or partially) belonging to the cylinders. This can be explained by looking at the filters’ output in the Supplementary Figures S6-S7, RORPO filter output is more uniform across the whole cylinders that it detects, and in consequence the segmentation by thresholding will capture better the whole shape of the object. The Frangi filter, on the other hand, was better than RORPO in detecting the inner voxels of the cylinders rather than the boundary, which is rather confounded by partial volume effects. From these results we can confidently accept that a PVS grows in diameter and length if the change is at least twice the maximum standard deviation of the diameter regardless of the filtering method used. The maximum standard deviation of the diameter for the Frangi filter was 1.65 mm and for the RORPO filter it was 0.79 mm. RORPO lower standard deviation can be explained by the uniformity of the filter output throughout all diameters (Supplementary Figure S9). Length measurements from using the Frangi filter were closer to the ideal segmentations (i.e., theoretical and considering voxels with at least 30% inside the cylinder) throughout the whole range analysed: 2 mm to 10 mm.

**Figure 10.**
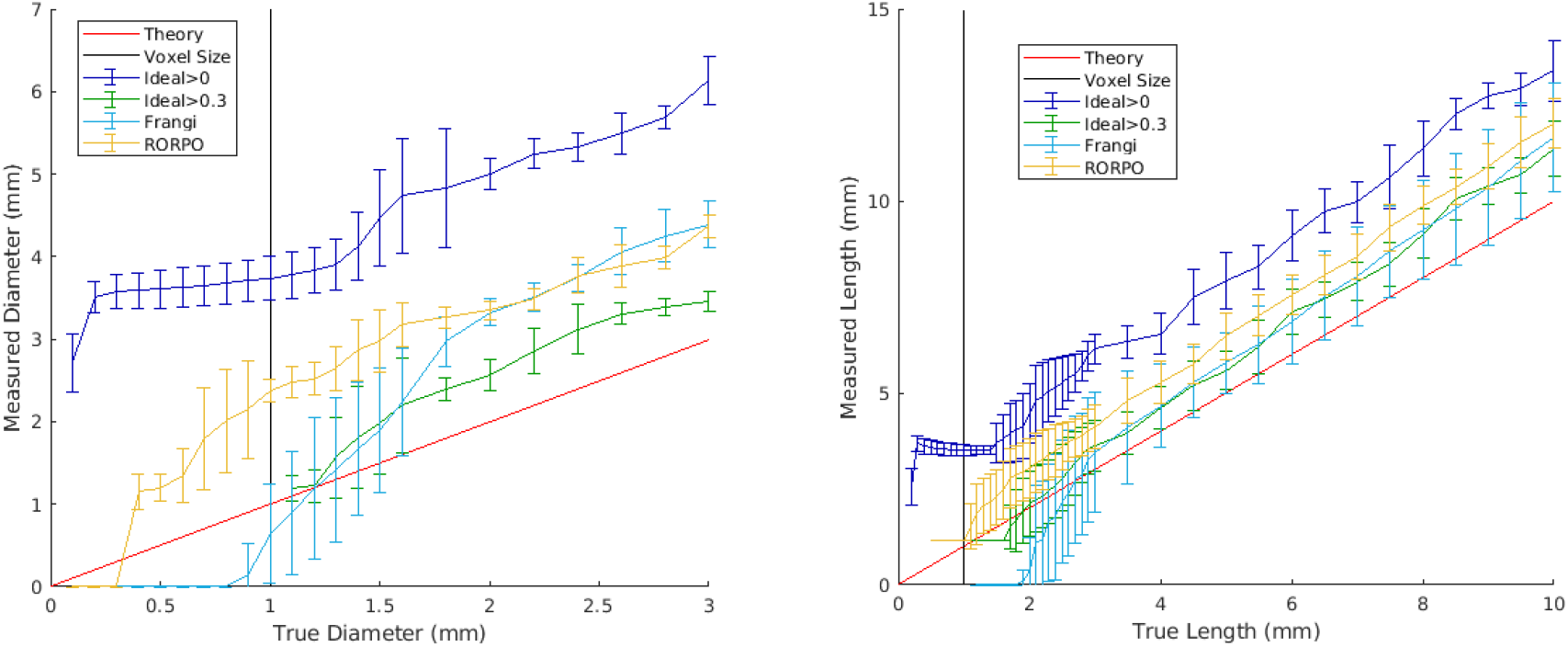
Comparison of the computed diameters and lengths for both filters for the *in-silico* phantom of isotropic voxels of 1 mm^3^. (Left) Diameters (Right) Lengths. The red line corresponds to the theoretical diameters, the black line corresonds to the voxel size of the scans, the dark blue line corresponds to the ideal segmentation of the *in-silico* phantom considering voxel with any proportion of the cylinder, the green line corresponds to the ideal segmentation of the in-silico phantom considering voxels with at least 30% inside the cylinder, the light blue line corresponds to the Frangi filter results and the yellow line corresponds to the RORPO filter results.

## Discussion

We have developed and made publicly available an *in-silico* phantom using the PVS-DROs presented by Bernal et al. (2022), for testing and evaluating methods and MRI-acquisition protocols that allow to assess PVS-like small tubular structures. The use of the *in-silico* and physical phantoms presented here can provide useful information on the accuracy, robustness and detectability of structures that can be considered a proxy for PVS visible in MRI, not only for brain studies, but for the study of PVS in any other body organ. They can also be used to assess precision and performance of other PVS computational analysis methods. The PVS-DRO model as used in our *in-silico* phantom can be tuned to agree with the spin-echo T2-weighted images of the UMC Utrecht physical phantom scanned in a 7T Phillips scanner with different voxel sizes and different orientations. Both phantoms represent the tubular shape of PVS using cylinders for easiness in their representation and manipulation. A more sophisticated simulation can be achieved using equation (3) for the high-resolved image, and using the non-uniform Fourier transform to sample the *k*-space of the desired scan for simulating the low-resolution image. This would allow also to simulate more complex scenarios such as Rician-noise and ghosting, as per Bernal et al. (2022). We could define curved PVS using a parametric equation of the curved line and generating the PVS binary mask by setting to 1 all voxels that are inside the given radius of the PVS. However, such a representation would have introduced ambiguities in the interpretation of the results, as in such scenario a single “PVS” would had different orientations across its dimensionality.

The results presented show that in all cases, regardless of the filtering or image processing methods used, the boundaries of the PVS representation are consistently overestimated due to a) the fact that these structures do not cross the voxels by their centre (Supplementary Table 1) and b) partial volume effect (as seen in Figure 3; and in Bernal et al.,2022). Therefore, as with many other MRI measures (Gassenmaier et al., 2020; Keenan et al., 2021; Illán-Gala et al., 2022; Nousiainen & Mäkelä, 2020), PVS measures from MRI are only a proxy of the real PVS burden and individual dimensions, which depend on image resolution and processing methods, and which needs to be considered when comparing PVS measurements between scans.

The success in the use of the Frangi filter in detecting PVS-like structures from MRI relies on a careful tuning of several parameters. The combination of parameters proposed by Ballerini et al. (2016, 2018) α=0.5, β=0.5 and c=500 (although c may change depending on the signal intensity of the input image), proved to yield the best results in all the simulations done so far (i.e., here, and in Bernal et al., 2022), while the recommended scaling range and ratios may vary depending on the spatial resolution (i.e., voxel size) of the image and intensity contrast. RORPO, on the contrary, does not have these requirements, and compared with Frangi, allows detecting smaller cylinders in their entirety more accurately. The segmentation of the cylinders using the Frangi filter, however, seems to be best suited for voxel sizes equal or larger than 0.4 mm-isotropic and cylinders larger than 1 mm diameter and 2 mm length. Hence, although the use of the RORPO filter may seem to outperform that of the Frangi filter in most of the discussed scenarios, to establish either filter as a better choice, we need to analyse other factors such as voxel anisotropy, presence of Gibbs artefacts, Rician noise, curved PVS, presence of other imaging confounds as lacunes and white matter hyperintensities, and grouping/clustering of PVS. Also, given that the PVS that are of interest clinically have larger sizes than the minimal dimensions for which Frangi seems to be best suited, in absence of artefacts or any confounding effects, the decision of whether to use one filtering method or the other comes down to the performance of the filter in the presence of these other factors.

The phantoms have limitations in terms of their representation of the living brain. Importantly, they do not include the white matter hyperintensties, lacunes, varied background signal in normal-appearing white matter, etc., that are commonly associated with having more PVS. Therefore, a more realistic *in-silico* phantom mimicking spatial interaction of the tubular structures with other lesion-like features representatives of white matter hyperintensities, strokes and lacunes, will be needed to ascertain the influence of these confounds in the segmentation of PVS-like structures and evaluate the capability of the image–processing methods to avoid false positives. While the discussed filters are designed to avoid objects that are not tube-like (e.g. blob-like and plate-like objects), the limits of these features needs to be further explored.

Although deep learning methods are still to be established for PVS-segmentations, promising results of experimental schemes have been emerging (Pham et al., 2022; Barisano et al., 2022). Further evaluation of these architectures using the phantoms presented here will hopefully help in the standardisation of the PVS segmentation with views at cross-studies analyses to advance the knowledge about the role of these structures in brain health.

## Acknowledgements

This study is mainly funded by the Hilary and Galen Weston Foundation under the Novel Biomarkers 2019 scheme (ref UB190097) administered by the Weston Brain Institute. This study received also funds from the Fondation Leducq Network of Excellence for the Study of Perivascular Spaces in Small Vessel Disease (16 CVD 05); the MRC Doctoral Training Programme in Precision Medicine (JB); the Row Fogo Charitable Trust (MCVH, FC) (BRO-D.FID3668413); and the UK Dementia Research Institute at the University of Edinburgh funded by the Medical Research Council, Alzheimer’s Society and Alzheimer’s Research UK (JMW).

## References

Aribisala, B.S., Riha, R.L., Valdes Hernandez, M., Muñoz Maniega, S., Cox, S., Radakovic, R., Taylor, A., Pattie, A., Corley, J., Redmond, P., Bastin, M.E., Starr, J., Deary, I., Wardlaw, J.M., 2020. Sleep and brain morphological changes in the eighth decade of life. Sleep Med. 65,152–158. https://doi.org/10.1016/j.sleep.2019.07.015

Aribisala, B.S., Valdés Hernández, M.D.C., Okely, J.A., Cox, S.R., Ballerini, L., Dickie, D.A., Wiseman, S.J., Riha, R.L., Muñoz Maniega, S., Radakovic, R., Taylor, A., Pattie, A., Corley, J., Redmond, P., Bastin, M.E., Deary, I., Wardlaw, J.M., 2023. Sleep quality, perivascular spaces and brain health markers in ageing - A longitudinal study in the Lothian Birth Cohort 1936. Sleep Med. 106, 123–131. https://doi.org/10.1016/j.sleep.2023.03.016

Aribisala, B.S., Wiseman, S., Morris, Z., Valdés-Hernández, M.C., Royle, N.A., Maniega, S.M., Gow, A.J., Corley, J., Bastin, M.E., Starr, J., Deary, I.J., Wardlaw, J.M., 2014. Circulating inflammatory markers are associated with magnetic resonance imaging-visible perivascular spaces but not directly with white matter hyperintensities. Stroke. 45(2),605–7. https://doi.org/10.1161/STROKEAHA.113.004059

Ballerini, L., Lovreglio, R., Valdés Hernández, M.D.C., Ramirez, J., MacIntosh, B.J., Black, S.E., Wardlaw, J.M., 2018. Perivascular Spaces Segmentation in Brain MRI Using Optimal 3D Filtering. Sci. Rep. 8, 1–11. https://doi.org/10.1038/s41598-018-19781-5

Ballerini, L., Lovreglio, R., Valdés Hernández, M.D.C., Gonzalez-Castro, V., Muñoz Maniega, S., Pellegrini, E., Bastin, M.E., Deary, I.J., Wardlaw, J.M., 2016. Application of the Ordered Logit Model to Optimising Frangi Filter Parameters for Segmentation of Perivascular Spaces. Procedia Computer Science 90, 61–67, ISSN 1877-0509, https://doi.org/10.1016/j.procs.2016.07.011

Baril, A.A., Pinheiro, A.A., Himali, J.J., Beiser, A., Sanchez, E., Pase, M.P., Seshadri, S., Demissie, S., Romero, J.R., 2022. Lighter sleep is associated with higher enlarged perivascular spaces burden in middle-aged and elderly individuals. Sleep Med. 100, 558–564. https://doi.org/10.1016/j.sleep.2022.10.006

Barisano, G., Lynch, K.M., Sibilia, F., Lan, H., Shih, N.C., Sepehrband, F., Choupan, J., 2022. Imaging perivascular space structure and function using brain MRI. Neuroimage. 257:119329. https://doi.org/10.1016/j.neuroimage.2022.119329

Berezuk, C., Ramirez, J., Gao, F., Scott, C.J., Huroy, M., Swartz, R.H., Murray, B.J., Black, S.E., Boulos, M.I., 2015. Virchow-Robin Spaces: Correlations with Polysomnography-Derived Sleep Parameters. Sleep. 38(6):853–8. https://doi.org/10.5665/sleep.4726

Bernal, J., Valdés-Hernández, M.D.C., Escudero, J., Duarte, R., Ballerini, L., Bastin, M.E., Deary, I.J., Thrippleton, M.J., Touyz, R.M., Wardlaw, J.M., 2022. Assessment of perivascular space filtering methods using a three-dimensional computational model. Magn Reson Imaging. 93, 33–51. https://doi.org/10.1016/j.mri.2022.07.016

Bouvy, W.H., Biessels, G.J., Kuijf, H.J., Kappelle, L.J., Luijten, P.R., Zwanenburg, J.J., 2014. Visualization of perivascular spaces and perforating arteries with 7 T magnetic resonance imaging. Invest Radiol.49(5), 307–13. https://doi.org/10.1097/RLI.0000000000000027

Del Brutto, O.H., Mera, R.M., Del Brutto, V.J., Castillo, P.R., 2019. Enlarged basal ganglia perivascular spaces and sleep parameters. A population-based study. Clin Neurol Neurosurg. 182:53–57. https://doi.org/10.1016/j.clineuro.2019.05.002

Donahue, E.K., Murdos, A., Jakowec, M.W., Sheikh-Bahaei, N., Toga, A.W., petzinger, G.M., Sepehrband, F., 2021. Global and regional changes in perivascular space in idiopathic and familial Parkinson’s disease. Mov. Disord. p. 28473, https://doi.org/10.1002/mds.28473

Duarte Coello, R.,,, Valdés Hernández, M.C.,,, Ballerini, L.,,, Bernal Moyano, J.,,, Chappell, F.M.,,, Brown, R.,,, Wardlaw, J.M.,,, Zwanenburg, J.,,, Van Der Velden, M.,,, Kuijf, H.J., 2023. Measurements of a perivascular spaces magnetic resonance imaging physical phantom and correspondent digital reference object model, 2023 [dataset]. University of Edinburgh. Centre for Clinical Brain Sciences. Department of Neuroimaging Sciences. https://doi.org/10.7488/ds/7454

Dubost, F., Yilmaz, P., Adams, H., Bortsova, G., Ikram, M.A., Niessen, W., Vernooij, M., de Bruijne, M., 2019. Enlarged perivascular spaces in brain MRI: Automated quantification in four regions. Neuroimage. 185:534–544. https://doi.org/10.1016/j.neuroimage.2018.10.026

Francis, F., Ballerini, L., Wardlaw, J.M., 2019. Perivascular spaces and their associations with risk factors, clinical disorders and neuroimaging features: A systematic review and meta-analysis. Int J Stroke. 14(4), 359–371. https://doi.org/10.1177/1747493019830321

Frangi, A.F., Niessen, W.J., Vincken, K.L., Viergever, M.A., 1998. Multiscale vessel enhancement filtering 130–137. https://doi.org/10.1007/BFb0056195

Gassenmaier, S., Tsiflikas, I., Maennlin, S. et al., 2020. Retrospective accuracy analysis of MRI based lesion size measurement in neuroblastic tumors: which sequence should we choose?. BMC Med Imaging 20, 105. https://doi.org/10.1186/s12880-020-00503-1

Hilal, S., Tan, C.S., Adams, H.H.H., Habes, M., Mok, V., Venketasubramanian, N., Hofer, E., Ikram, M.K., Abrigo, J., Vernooij, M.W., Chen, C., Hosten, N., Volzke, H., Grabe, H.J., Schmidt, R., Ikram, M.A., 2018. Enlarged perivascular spaces and cognition: A meta-analysis of 5 population-based studies. Neurology. 91(9), e832–e842. https://doi.org/10.1212/WNL.0000000000006079

Illán-Gala, I., Nigro, S., Vande Vrede, L., Falgàs, N., Heuer, H.W., Painous, C., Compta, Y., Martí, M.J., Montal, V., Pagonabarraga, J., Kulisevsky, J., Lleó, A., Fortea, J., Logroscino, G., Quattrone, A., Perry, D.C., Gorno-Tempini, M.L., Rosen, H.J., Grinberg, L.T., Spina, S., La Joie, R., Rabinovici, G.D., Miller, B.L., Rojas, J.C., Seeley, W.W., Boxer, A.L., 2022. Diagnostic Accuracy of Magnetic Resonance Imaging Measures of Brain Atrophy Across the Spectrum of Progressive Supranuclear Palsy and Corticobasal Degeneration. JAMA Netw Open. 5(4),p e229588. https://doi.org/10.1001/jamanetworkopen.2022.9588. Erratum in: JAMA Netw Open. 2022 May 2;5(5), e2217977.

Keenan, K.E., Gimbutas, Z., Dienstfrey, A., Stupic, K.F., Boss, M.A., Russek, S.E., Chenevert, T.L., Prasad, P.V., Guo, J., Reddick, W.E., Cecil, K.M., Shukla-Dave, A., Aramburu Nunez, D., Shridhar Konar, A., Liu, M.Z., Jambawalikar, S.R., Schwartz, L.H., Zheng, J., Hu, P., Jackson, E.F., 2021. Multi-site, multi-platform comparison of MRI T1 measurement using the system phantom. PLoS One. 16(6),p e0252966. https://doi.org/10.1371/journal.pone.0252966

Merveille, O., Talbot, H., Najman, L., Passat, N., 2018. Curvilinear Structure Analysis by Ranking the Orientation Responses of Path Operators. IEEE Trans. Pattern Anal. Mach. Intell. 40,304–317. https://doi.org/10.1109/TPAMI.2017.2672972

Nousiainen, K., Mäkelä, T., 2020. Measuring geometric accuracy in magnetic resonance imaging with 3D-printed phantom and nonrigid image registration. Magn Reson Mater Phy 33, 401–410. https://doi.org/10.1007/s10334-019-00788-6

Passiak, B.S., Liu, D., Kresge, H.A., Cambronero, F.E., Pechman, K.R., Osborn, K.E., Gifford, K.A., Hohman, T.J., Schrag, M.S., Davis, L.T., Jefferson, A.L., 2019. Perivascular spaces contribute to cognition beyond other small vessel disease markers. Neurology. 92(12), e1309–e1321. https://doi.org/10.1212/WNL.0000000000007124

Patankar, T.F., Mitra, D., Varma, A., Snowden, J., Neary, D., Jackson, A., 2005. Dilatation of the Virchow-Robin space is a sensitive indicator of cerebral microvascular disease: study in elderly patients with dementia. AJNR Am J Neuroradiol. 26(6),1512–1520.

Pham, W., Lynch, M., Spitz, G., O’Brien, T., Vivash, L., Sinclair, B., Law, M., 2022. A critical guide to the automated quantification of perivascular spaces in magnetic resonance imaging. Front. Neurosci. 16:1021311. https://doi.org/10.3389/fnins.2022.1021311

Potter, G.M., Doubal, F.N., Jackson, C.A., Chappell, F.M., Sudlow, C.L., Dennis, M.S., Wardlaw, J.M.,2015. Enlarged perivascular spaces and cerebral small vessel disease. Int J Stroke. 10(3),376–381. https://doi.org/10.1111/ijs.12054

Sepehrband, F., Barisano, G., Sheikh-Bahaei, N., Choupan, J., Cabeen, R.P., Lynch, K.M., Crawford, M.S., Lan, H., Mack, W.J., Chu,i H.C., Ringman, J.M., Toga, A.W., 2021. Alzheimer’s Disease Neuroimaging Initiative. Volumetric distribution of perivascular space in relation to mild cognitive impairment. Neurobiol Aging. 99:28–43. https://doi.org/10.1016/j.neurobiolaging.2020.12.010

Valdés Hernández, M. del C., Piper, R.J., Wang, X., Deary, I.J.,Wardlaw, J.M., 2013. Towards the automatic computational assessment of enlarged perivascular spaces on brain magnetic resonance images: A systematic review. J. Magn. Reson. Imaging. 38: 774–785. https://doi.org/10.1002/jmri.24047

Wardlaw, J.M., Smith, E.E., Biessels, G.J., Cordonnier, C., Fazekas, F., Frayne, R., Lindley, R.I., O’Brien, J.T., Barkhof, F., Benavente, O.R., Black, S.E., Brayne, C., Breteler, M., Chabriat, H., DeCarli, C., de Leeuw, F.E., Doubal, F., Duering, M., Fox, N.C., Greenberg, S., Hachinski, V., Kilimann, I., Mok, V., Oostenbrugge R. van Pantoni, L., Speck, O., Stephan, B.C.M., Teipel, S., Viswanathan, A., Werring, D., Chen, C., Smith, C., van Buchem, M., Norrving, B., Gorelick, P.B., Dichgans, M., 2013. Neuroimaging standards for research into small vessel disease and its contribution to ageing and neurodegeneration. Lancet Neurol. 12,822–838. https://doi.org/10.1016/S1474-4422(13)70124-8

Wardlaw, J.M., Benveniste, H., Nedergaard, M., Zlokovic, B.V., Mestre, H., Lee, H., Douba,l F.N., Brown, R., Ramirez, J., Macintosh, B.J., et al., 2020. Perivascular spaces in the brain: anatomy, physiology and pathology. Nature Reviews Neurology, 16, 137–153.

