## Supplementary for "On the detectability and accuracy of computational measurements of enlarged perivascular spaces from magnetic resonance images"

Supplementary material

Supplementary Table S1. Scan parameters of the 7T turbo-spin-echo (TSE) sequence used to obtain the images from the physical phantom

| Parameters |  |  |  |  |  |
| --- | --- | --- | --- | --- | --- |
| Acquired isotropic resolution (mm) | 1.0 | 0.9 | 0.8 | 0.7 | 0.6 |
| Reconstructed isotropic resolution^1^ | 0.5 | 0.45 | 0.4 | 0.35 | 0.3 |
| FOV (mm) | 150x176x50 | 150x177x50 | 150x178x50 | 150x150x50 | 150x150x50 |
| TSE factor | 176 | 176 | 178 | 178 | 177 |
| Echo spacing | 2.6 | 2.7 | 2.9 | 3.2 | 3.5 |
| Flip angle sweep (min, max1-max2)^2^ | 10, 70-90 | 10, 70-90 | 10, 70-90 | 10, 70-90 | 10, 70-90 |
| TR (ms) | 3000 | 3000 | 3000 | 3000 | 3000 |
| TE / TEeq (ms)^3^ | 240 / 42 | 254 / 44 | 272 / 46 | 297 / 50 | 326 / 55 |
| Bandwidth (Hz/pix) | 1354 | 1225 | 1089 | 953 | 830 |
| Scan duration (min:s)^4^ | 5:03 | 5:39 | 6:21 | 7:15 | 8:27 |

^1^ Reconstructed resolution: resolution of the reconstructed images, which is achieved by zero-filling the k-space before performing the Fourier transforms during the reconstruction; (vendor-supplied option in the protocol).

^2^ Refocusing flip angle sweep as described by Busse et al (2006)^Ɨ^: a tissue specific sweep leading to constant signal for a tissue type with specified T1/T2 until refocusing angle max1 is reached after which the refocusing flip angle linearly increases to max2. For this sweep the T1/T2 of white matter was used (1600/45ms).

^3^ TEeq: Equivalent echo time that yields similar contrast in a normal spin-echo sequence for the specified tissue as used in calculating the refocusing flip angle sweep.

^4^ No acceleration was used (i.e. SENSE factors 1.0)

^Ɨ^ Busse RF, Hariharan H, Vu A, Brittain JH. Fast spin echo sequences with very long echo trains: design of variable refocusing flip angle schedules and generation of clinical T2 contrast. Magn Reson Med. 2006 May; 55(5):1030-7. doi: 10.1002/mrm.20863. PMID: 16598719.

Supplementary Table S2 Diagonal and oblique lengths in isotropic voxels (from 0.5 to 1.0 mm-isotropic), depending on the spatial resolution of the scan.

| Setting (voxel size) | l2D (in-plane measurement in mm) | l3D (cross-plane measurement in mm) |
| --- | --- | --- |
| 0.5 x 0.5 x 0.5 mm | 0.71 | 0.87 |
| 0.6 x 0.6 x 0.6 mm | 0.85 | 1.03 |
| 0.7 x 0.7 x 0.7 mm | 0.99 | 1.21 |
| 0.8 x 0.8 x 0.8 mm | 1.13 | 1.39 |
| 0.9 x 0.9 x 0.9 mm | 1.27 | 1.56 |
| 1.0 x 1.0 x 1.0 mm | 1.41 | 1.73 |

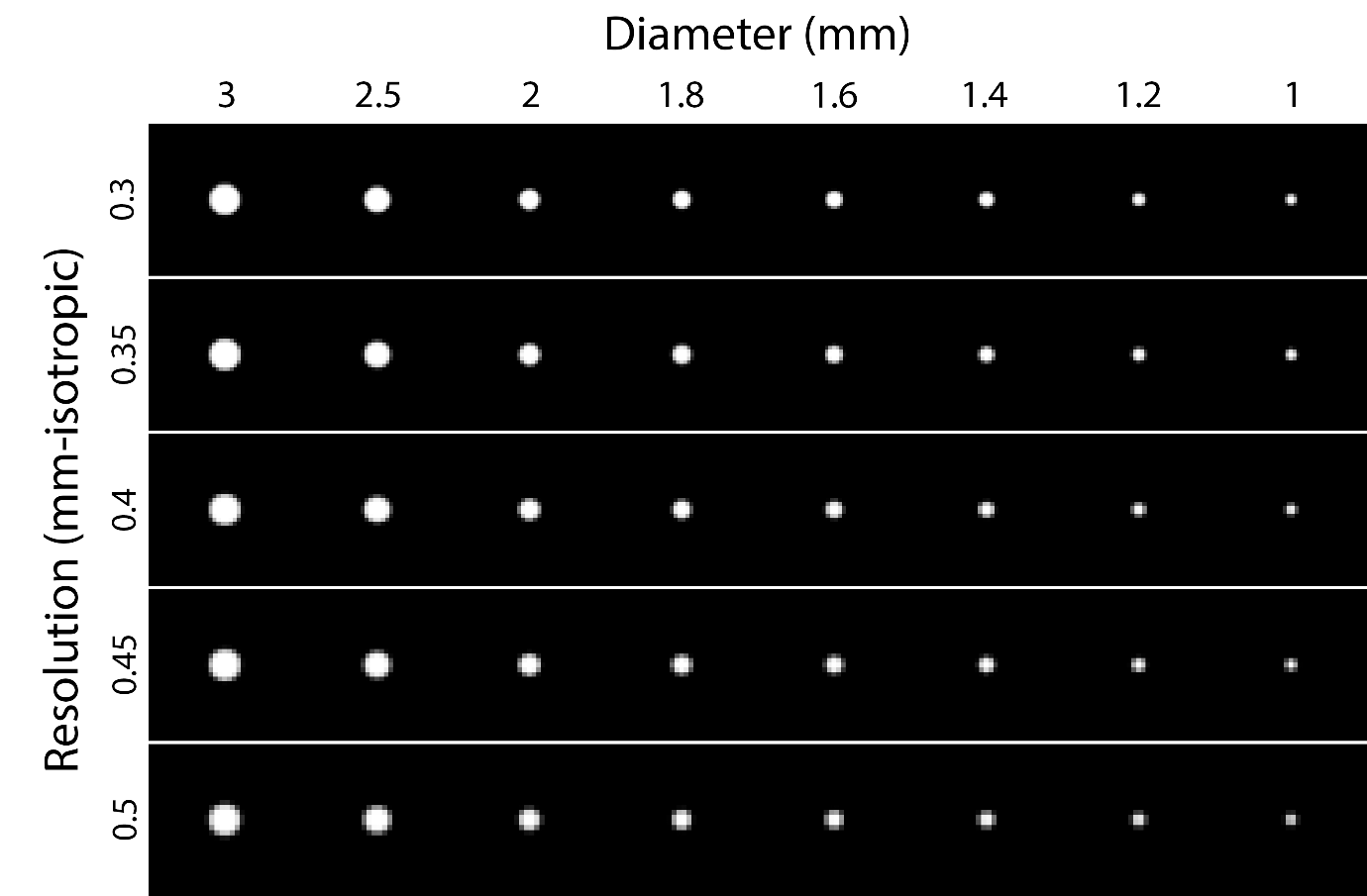

Supplementary Figure S1. Example of the simulated PVS diameters in the in-silico phantom. From top to bottom, voxel size 0.3, 0.35, 0.4, 0.45 and 0.5 cubic millimetres. From left to right, PVS diameters 3.0, 2.5, 2.0, 1.8, 1.6, 1.2, 1.0, and 0.5 millimetres.

Supplementary Table S3. Diameter (Ø) measurements in mm using the Frangi filter to identify the cylinders of the physical phantom. Average of the measurements of the three holes with equal diameter (Ave Ø), and total range of variation among the three measurements (Maximum Ø – Minimum Ø) for each voxel size (VS, in mm-isotropic).

| Real  Ø | VS 0.3 | | VS 0.35 | | VS 0.4 | | VS 0.45 | | VS 0.5 | |
| --- | --- | --- | --- | --- | --- | --- | --- | --- | --- | --- |
|  | Ave Ø | Max-Min | Ave Ø | Max-Min | Ave Ø | Max-Min | Ave Ø | Max-Min | Ave Ø | Max-Min |
| 0.2 | 0 | 0 | 0 | 0 | 0 | 0 | 0 | 0 | 0 | 0 |
| 0.3 | 0 | 0 | 0 | 0 | 0 | 0 | 0 | 0 | 0 | 0 |
| 0.4 | 0 | 0 | 0 | 0 | 0 | 0 | 0 | 0 | 0 | 0 |
| 0.5 | 0.11 | 0.33 | 0 | 0 | 0 | 0 | 0 | 0 | 0 | 0 |
| 0.6 | 1.095 | 0.059 | 1.013 | 0.47 | 0.30 | 0.45 | 0 | 0 | 0 | 0 |
| 0.7 | 1.27 | 0.16 | 1.40 | 0.16 | 1.51 | 0.11 | 0 | 0 | 0 | 0 |
| 0.8 | 1.71 | 0.040 | 1.61 | 0.039 | 1.73 | 0.073 | 0.17 | 0.52 | 0 | 0 |
| 0.9 | 1.96 | 0.025 | 1.92 | 0.18 | 1.76 | 0.24 | 1.56 | 0.69 | 0.38 | 1.13 |
| 1 | 2.18 | 0.092 | 2.27 | 0.14 | 2.047 | 0.029 | 1.91 | 0.34 | 1.74 | 0.94 |
| 1.2 | 2.44 | 0.14 | 2.17 | 0.15 | 2.49 | 0.064 | 2.42 | 0.18 | 2.038 | 0.21 |
| 1.4 | 2.63 | 0.16 | 2.46 | 0.15 | 2.71 | 0.047 | 2.66 | 0.26 | 2.36 | 0.25 |
| 1.6 | 2.72 | 0.13 | 2.86 | 0.17 | 2.88 | 0.17 | 2.85 | 0.24 | 2.48 | 0.12 |
| 1.8 | 2.88 | 0.14 | 3.056 | 0.066 | 3.074 | 0.13 | 3.073 | 0.17 | 2.74 | 0.32 |
| 2 | 2.92 | 0.11 | 3.044 | 0.22 | 3.12 | 0.15 | 3.12 | 0.16 | 2.81 | 0.40 |
| 2.5 | 3.062 | 0.13 | 3.16 | 0.18 | 3.27 | 0.24 | 3.25 | 0.24 | 2.95 | 0.25 |
| 3 | 3.42 | 0.11 | 3.32 | 0.27 | 3.62 | 0.074 | 3.68 | 0.15 | 3.35 | 0.22 |

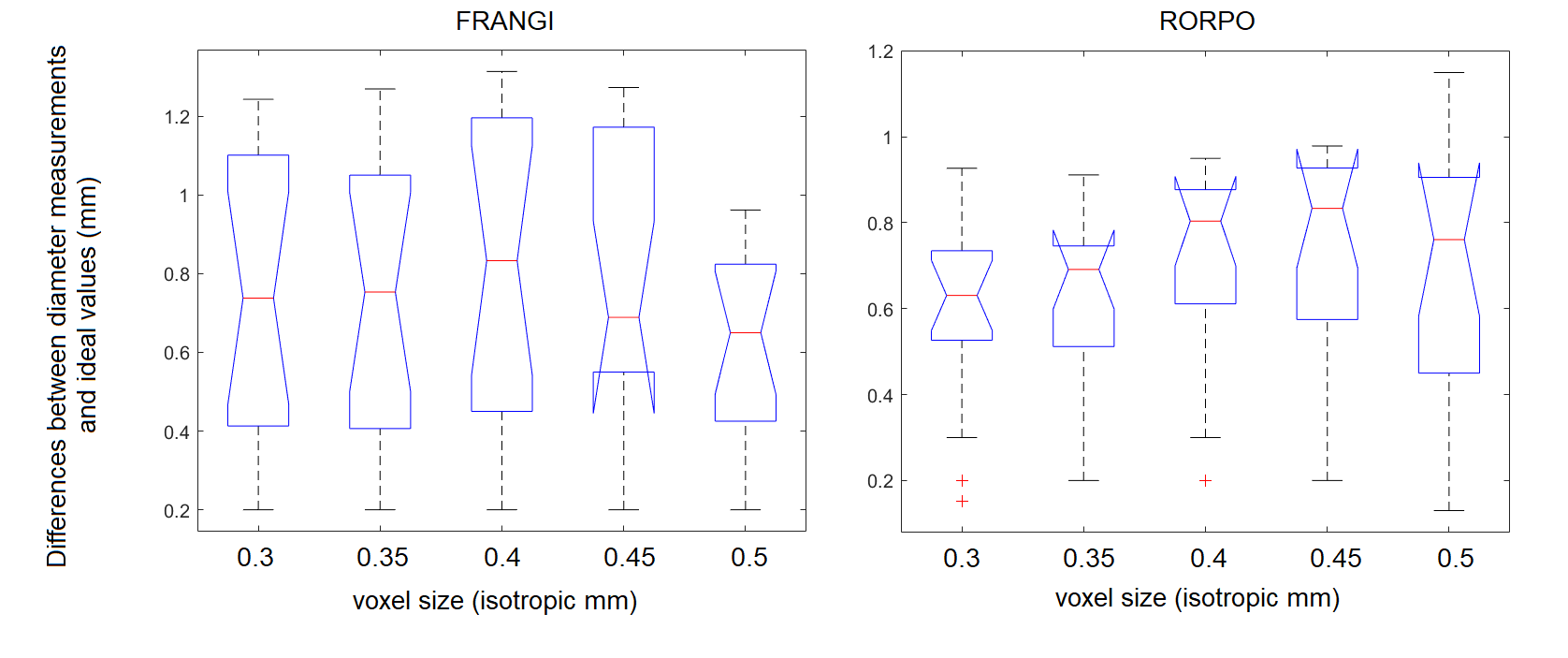

Supplementary Figure S2. Box plots of the absolute differences between diameter measurements and ideal values for different voxel sizes as measured using the Frangi (left hand side) and RORPO (right hand side) filters.

Supplementary Table S4. Results of the statistical test comparing the average measurements of the three cylinders with equal diameter, range of variation of the three measurements per diameter, and absolute differences of the average measurements with respect to the ideal segmentation, using RORPO and Frangi filtering methods. h = 0 indicates a failure to reject the null hypothesis and p-values (p) are given.

| **Statistical test** | **Voxel size** | **0.3 mm-isotropic** | **0.4 mm-isotropic** | **0.5 mm-isotropic** |
| --- | --- | --- | --- | --- |
| **Wilcoxon sign rank:**  paired, two-sided test for the null hypothesis that the data from RORPO – Frangi comes from a distribution with zero median | Average measurements | p = 0.36, h = 0 | p = 0.50, h =0 | **p = 0.009, h = 1** |
|  | Range of variation | p = 0.62, h = 0 | p = 0.91, h = 0 | p = 0.52, h = 0 |
|  | Absolute differences with the ideal value | p = 0.068, h = 0 | p = 0.17, h = 0 | p = 0.34, h = 0 |

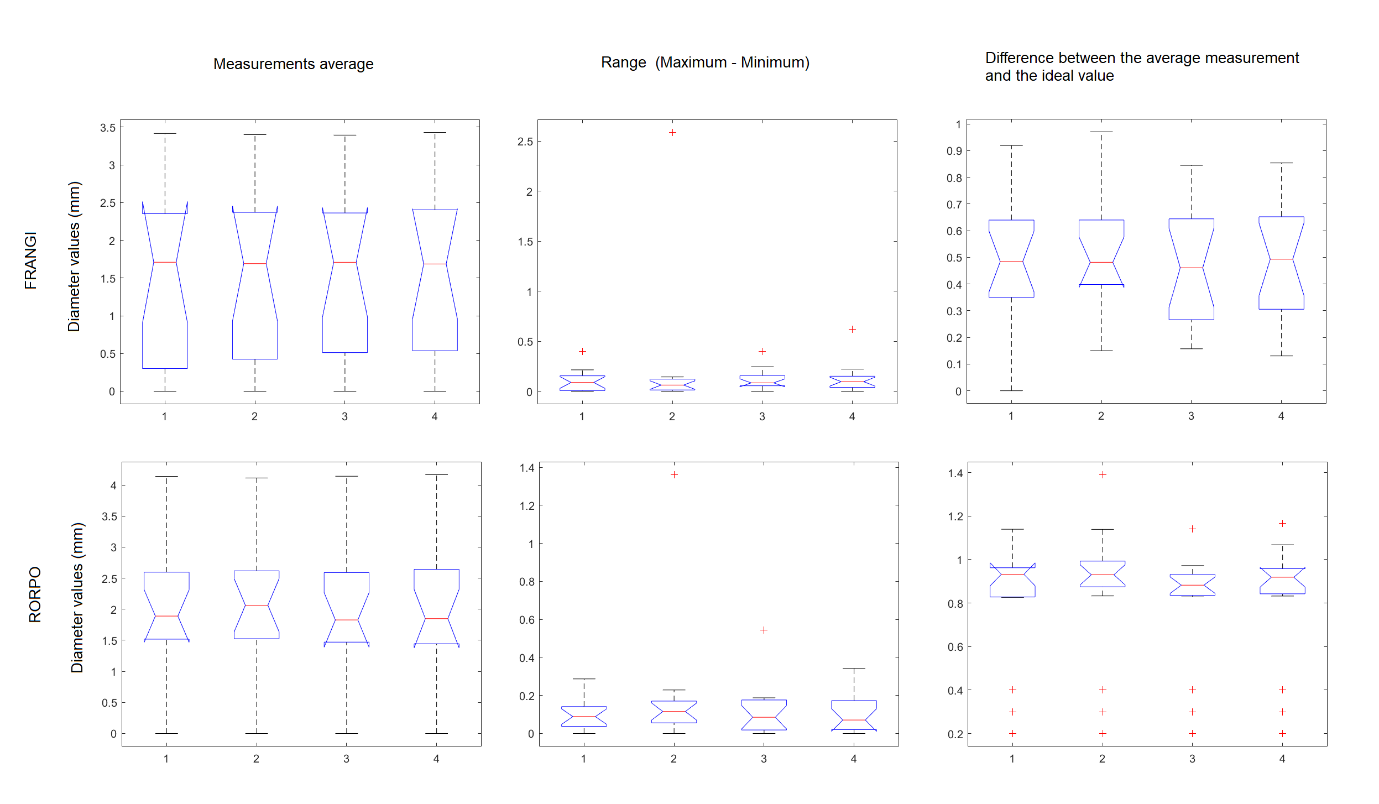

Supplementary Figure S3. Box plots comparing: a) the average measurements of the three cylinders with equal diameter (left hand side column) acquired under four different angles, b) range of variation of the three measurements per diameter (middle column) acquired under four different angles, and c) absolute differences of the average measurements with respect to the ideal segmentation (right hand side column), using RORPO (row below) and Frangi (row above) filtering methods. Numbers from 1 to 4 in the horizontal axes of each plot correspond to orientations: coronal, 15^o^, 30^o^ and 45^o^ around feet-head axis with respect to the coronal, respectively.

Supplementary Table S5. Mean and Standard Deviation (SD) of the differences between the valid measurements (i.e., excluding those cylinders that were undetected) and ideal cylinder diameters, from Bland-Altman analyses (Figure 7 in manuscript)

| Voxel size (VS) | Mean difference Frangi (mm) | SD Frangi (mm) | Mean difference RORPO (mm) | SD RORPO (mm) |
| --- | --- | --- | --- | --- |
| 0.3 mm-isotropic | 0.87 | 0.34 | 0.68 | 0.14 |
| 0.35 mm-isotropic | 0.90 | 0.32 | 0.71 | 0.13 |
| 0.4 mm-isotropic | 0.96 | 0.36 | 0.81 | 0.13 |
| 0.45 mm-isotropic | 0.97 | 0.37 | 0.85 | 0.14 |
| 0.5 mm-isotropic | 0.72 | 0.30 | 0.84 | 0.21 |

Legend: Differences are calculated as ideal value – measured value

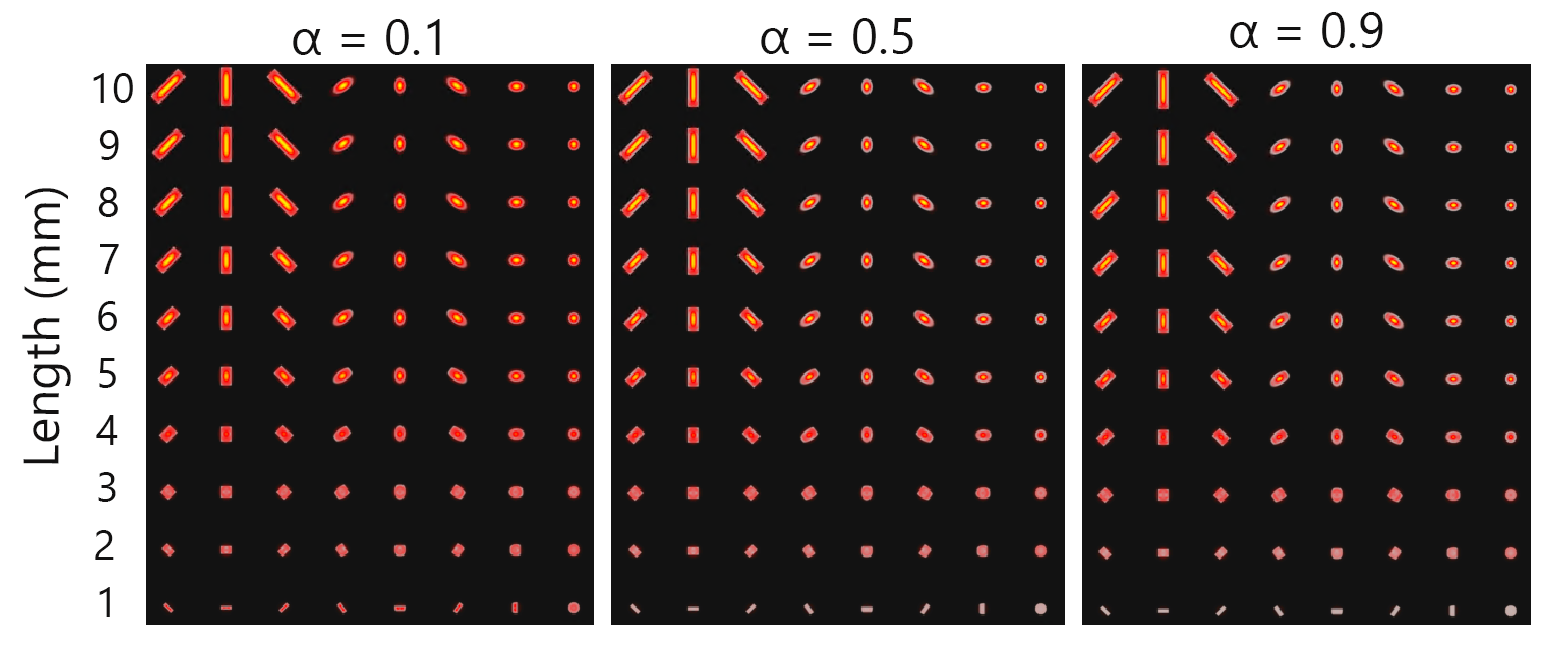

Supplementary Figure S4. Impact of Frangi Filter’s α parameter in the *in-silico* cylinders at 0.3 mm-isotropic voxels. The cylinders’ lengths from bottom to top are 1 to 10 mm in increments of 1mm. From left to right panels, α is 0.1, 0.5 and 0.9. The parameter α controls the sensitivity of the dissimilarity measures between tube-like and plate-like structures. Values of α that are too low, allow more plate-like structures to be highlighted.

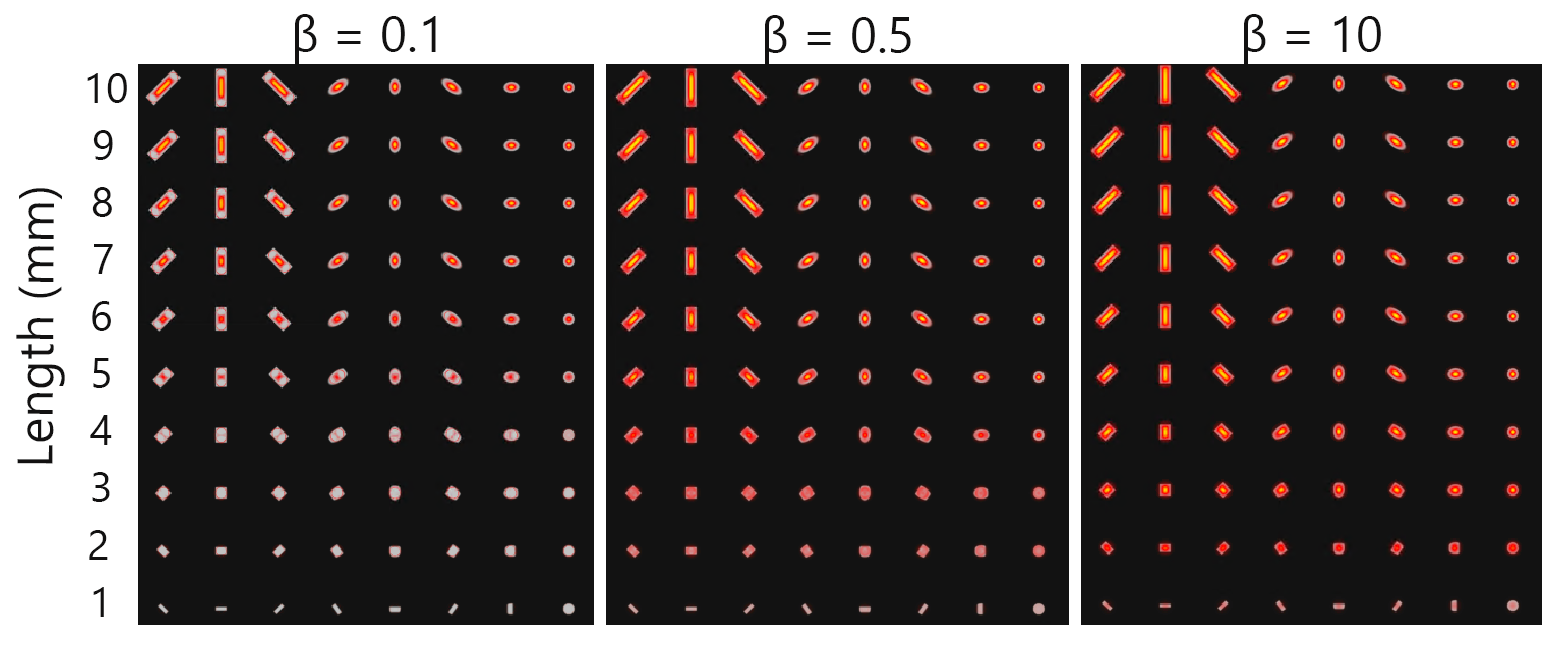

Supplementary Figure S5. Impact of Frangi Filter’s β parameter in the *in-silico* cylinders at 0.3 mm-isotropic voxels. The lengths from the cylinders from bottom to top are of 1 to 10 mm in increments of 1mm. From left to right, β is 0.1, 0.5 and 10. The β parameter controls the sensitivity of the dissimilarity measures between tube-like and blob-like structures. Lower values of β reduce the sensitivity of short cylinders and the filter response in the extremes of the cylinder. Values of β that are too high, allow more blob-like structures to be highlighted.

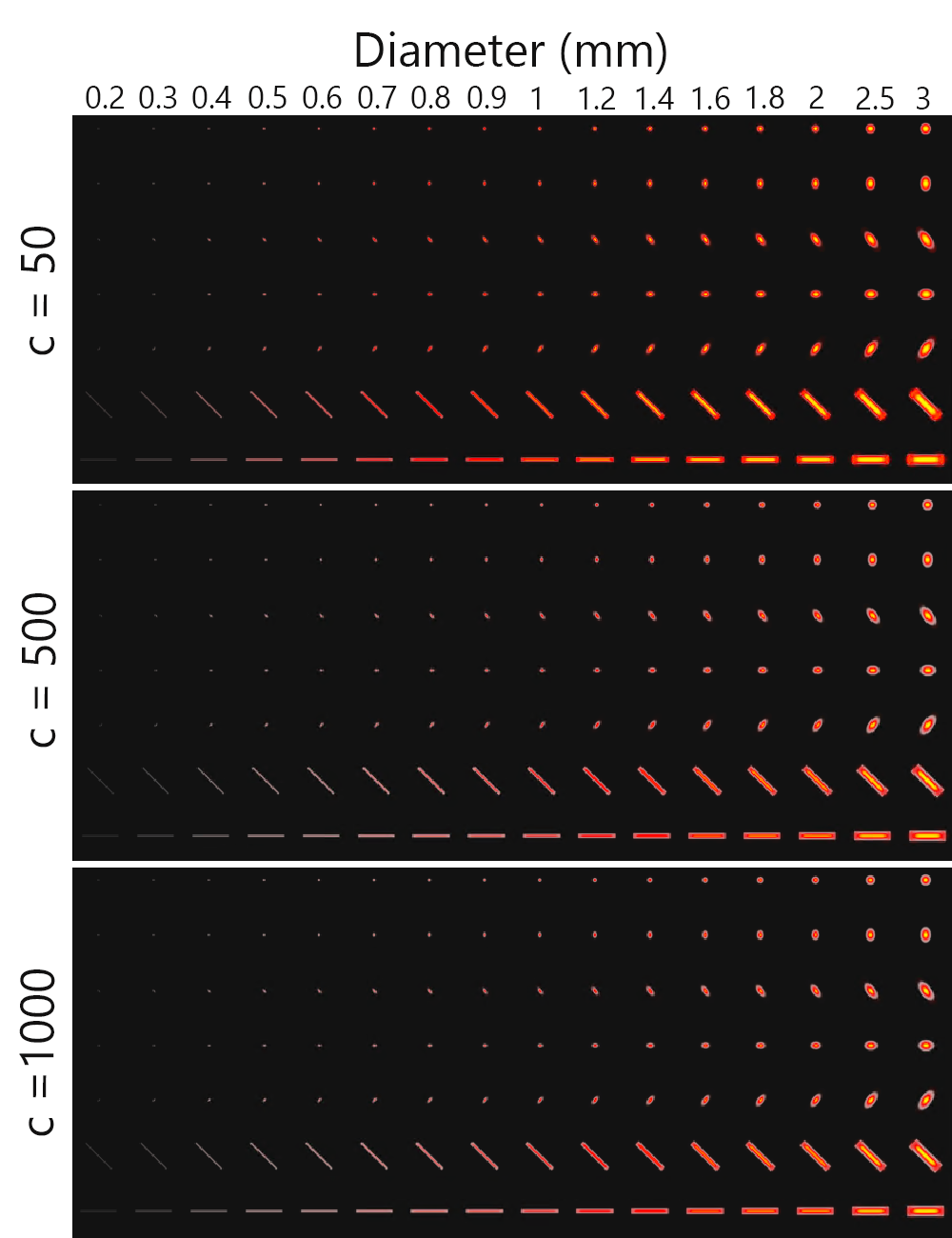

Supplementary Figure S6. Impact of the parameter c of the Frangi Filter in the *in-silico* phantom at 0.3 mm-isotropic voxels. The diameter from the cylinders from left to right are 0.2, 0.3, 0.4, 0.5, 0.6, 0.7, 0.8, 0.9, 1, 1.2, 1.4, 1.6, 1.8, 2, 2.5 and 3 mm. From top to bottom, c is 50, 500 and 100. The c parameter controls the sensitivity of the signal intensity of the object. In this case, the maximum signal intensity is 255. Lower values of c increase the sensitivity of capturing the objects but could also increase false positives.

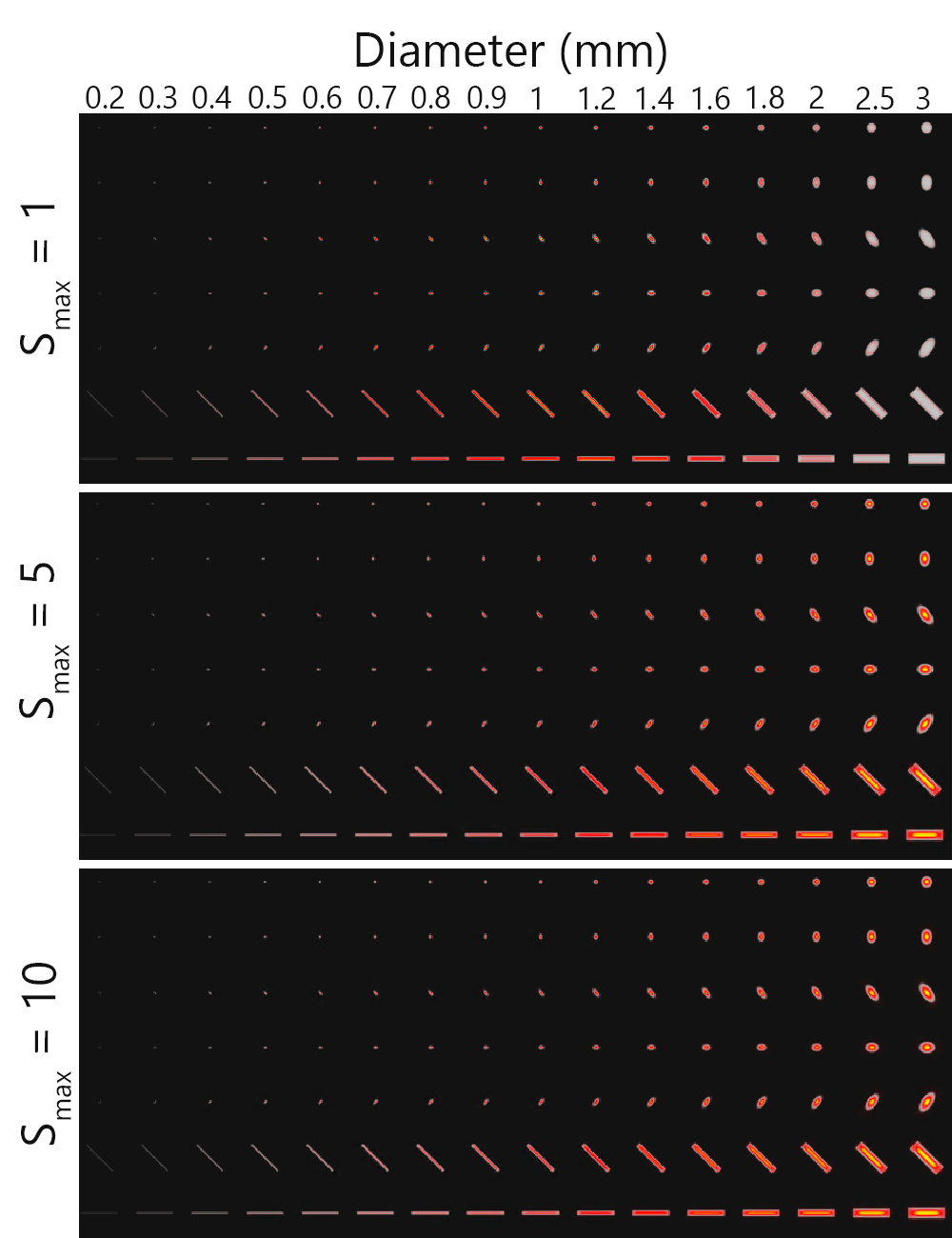

Supplementary Figure S7. Impact of the maximum scale parameter of the Frangi Filter in the *in-silico* phantom at 0.3 mm-isotropic voxels. The diameter from the cylinders from left to right are 0.2, 0.3, 0.4, 0.5, 0.6, 0.7, 0.8, 0.9, 1, 1.2, 1.4, 1.6, 1.8, 2, 2.5 and 3mm. From top to bottom, the maximum scale is 1, 5 and 10. The maximum scale parameter controls the sensitivity to the size of the cylinders. Higher values of the maximum scale will detect bigger tubular objects.

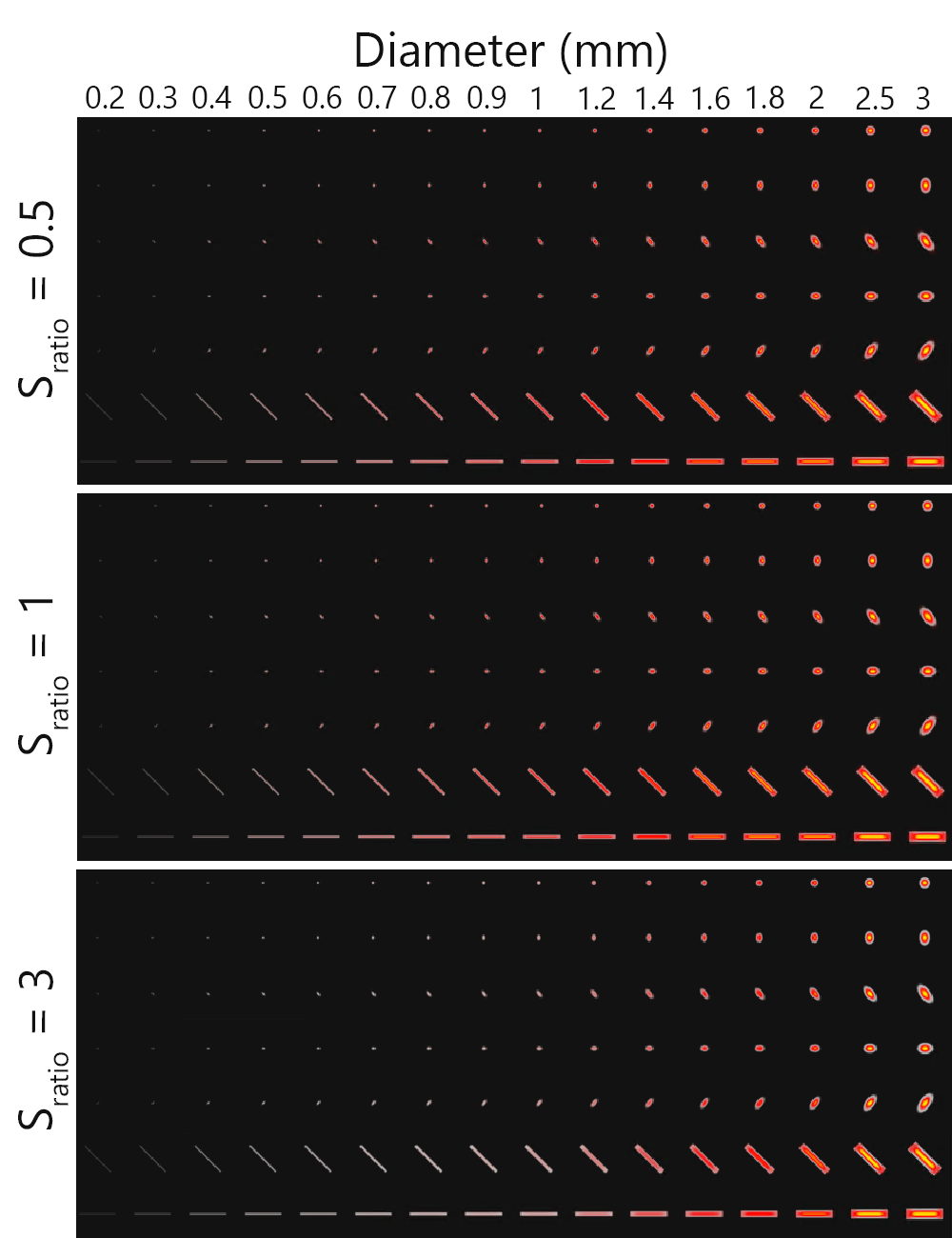

Supplementary Figure S8. Impact of the scale ratio of the Frangi Filter in the *in-silico* phantom with 0.3 mm-isotropic voxels. The diameter from the cylinders from left to right are 0.2, 0.3, 0.4, 0.5, 0.6, 0.7, 0.8, 0.9, 1, 1.2, 1.4, 1.6, 1.8, 2, 2.5 and 3mm. From top to bottom, the scales were evaluated every 0.15, 0.3 and 0.9 mm. By evaluating finer scales, the output of the filter will detect better the structures but increase the computational time. By evaluating the scales too coarse, the computational time can be reduced but some cylinders may not be detected correctly.

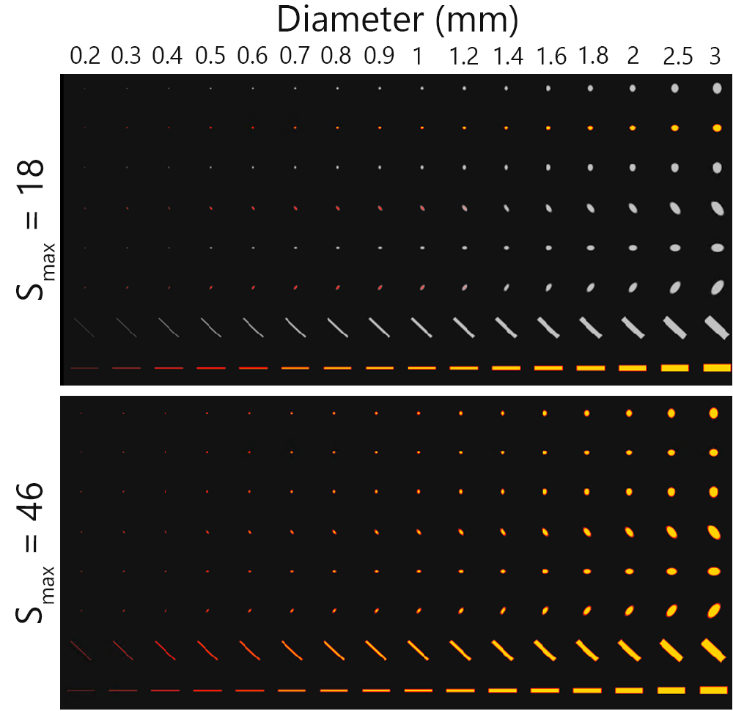

Supplementary Figure S9. Impact of the maximum scale parameter of the RORPO Filter in the *in-silico* phantom at 0.3 mm-isotropic voxels. The diameter from the cylinders from left to right are 0.2, 0.3, 0.4, 0.5, 0.6, 0.7, 0.8, 0.9, 1, 1.2, 1.4, 1.6, 1.8, 2, 2.5 and 3mm. From top to bottom, the maximum scale is 18 and 46. The maximum scale parameter controls the sensitivity to the size of the cylinders and has a bigger effect on objects that are not aligned to the Cartesian coordinates. Higher values of the maximum scale will detect bigger tubular objects.
